# Pan-Canadian survey on the impact of the COVID-19 pandemic on cervical cancer screening and management

**DOI:** 10.1101/2022.09.23.22279458

**Authors:** Mariam El-Zein, Rami Ali, Eliya Farah, Sarah Botting-Provost, Eduardo L. Franco, Survey Study Group

## Abstract

**Background:** The COVID-19 pandemic has caused disruptions to cancer care by delaying diagnoses and treatment, presenting challenges and uncertainties for both patients and physicians. We conducted a nationwide online survey to investigate the effects of the pandemic and capture modifications, prompted by pandemic-related control measures, on cervical cancer screening-related activities from mid-March to mid-August 2020, across Canada.

**Methods:** The survey consisted of 61 questions related to the continuum of care in cervical cancer screening and treatment: appointment scheduling, tests, colposcopy, follow-up, treatment of pre-cancerous lesions/cancer, and telemedicine. We piloted the survey with 21 Canadian experts in cervical cancer prevention and care. We partnered with the Society of Canadian Colposcopists, Society of Gynecologic Oncology of Canada, Canadian Association of Pathologists, and Society of Obstetricians and Gynecologists of Canada, which distributed the survey to their members via email. We reached out to family physicians and nurse practitioners via MDBriefCase. The survey was also posted on McGill Channels (Department of Family Medicine News and Events) and social media platforms. The data were analyzed descriptively.

**Results:** Unique responses were collected from 510 participants (16 November 2020 - 28 February 2021), representing 418 fully- and 92 partially-completed surveys. Responses were from Ontario (41.0%), British Columbia (21.0%), and Alberta (12.8%), and mostly comprised family physicians/general practitioners (43.7%), and gynecologist/obstetrician professionals (21.6%). Cancelled screening appointments were mainly reported by family physicians/general practitioners (28.3%), followed by gynecologist/obstetrician professionals (19.8%), and primarily occurred in private clinics (30.5%). Decreases in the number of screening Pap tests and colposcopy procedures were consistently observed across Canadian provinces. About 90% reported that their practice/institution adopted telemedicine to communicate with patients.

**Conclusions:** The area most severely impacted by the pandemic was appointment scheduling, with an important level of cancellations reported. Survey results may inform resumptions of various fronts in cervical cancer screening and management.

## Introduction

Following the announcement of the coronavirus disease 2019 (COVID-19) pandemic by the World Health Organization in mid-March 2020,^1^ the severe acute respiratory syndrome coronavirus 2 (SARS-CoV-2) has since spread across the globe, resulting in enough severe illness to overwhelm the healthcare system.^2,3^ The immediate, preventive measures taken in response have adversely affected an entire range of activities, specifically those related to cancer control, prevention, and care.^4^ As a result, cancer screening and treatment services have been scaled back to conserve resources, increase capacity for managing COVID-19 patients, and lower the risk of infections among cancer patients worldwide, particularly during full and partial lockdown periods. Elective surgery, chemotherapy, and radiotherapy procedures have been postponed or modified, which necessitated rapid adaptation by the medical community and adjustment in health services, including the use of telehealth. About one third of family medicine physicians in North America reported delaying cancer screening in the early phase of the pandemic and while some physicians reported high use of telehealth, most reported that reductions in cancer screenings would lead to increased incidence of late-stage cancers.^5^

Significant decreases in cancer diagnoses have been reported in multiple affected countries,^6,7^ alongside challenges in delivering timely cancer care to patients.^8–11^ According to a survey carried out between May 22 and June 10, 2020 by the Canadian Cancer Survivor Network, 54% of patients reported that their cancer care-related appointments were cancelled, postponed, or rescheduled due to the pandemic, and 71% expressed concerns about their ability to receive proper care, testing, and follow-up appointments in a timely fashion.^12^ A stochastic microsimulation model using data from the Canadian Cancer Registry predicted that pandemic-related cancer care disruptions (March 2020 - Jun 2021) could lead to 21,247 (2.0%) more cancer deaths in Canada between 2020 and 2030, if treatment capacity in 2021 were to recover to 2019 pre-pandemic levels. ^13^

Breast and cervical cancer screening tests in the United States declined by 87% and 84%, respectively, during April 2020 compared with the previous 5-year averages for the month of April.^14^ In Ontario, Canada, there were 41% fewer screening tests delivered in 2020 for breast, cervical, colorectal and lung cancer than in 2019.^15^ A population-based study in Ontario, Canada found that, between March and August 2020, the average monthly number of cytology tests, colposcopies, and treatments decreased by 63.8%, 39.7%, and 31.1%, respectively, compared to the same months in 2019 and that on average there were 292 fewer high-grade cytological abnormalities (decrease by 51.0%) detected each month.^16^

We conducted a national cross-sectional survey-based descriptive study among healthcare professionals to 1) assess the early impact of the pandemic on cervical cancer screening, diagnosis, management, and treatment services across Canada and 2) identify actionable approaches used by experts to mitigate the impact of the pandemic on their practice.

## Materials and Methods

### Target population

The survey questions were formulated to gather the opinions and firsthand experiences of colposcopists, colposcopy registered nurses, registered practical nurses, cytopathologists, technologists, general practitioners, family physicians, obstetrician and gynecology staff, gynecological oncologists, gynecology nurses, pathologists, and physician assistants working in private and public health institutions in Canada.

### Survey design, development, and validation

We used the Checklist for Reporting Results of Internet E-Surveys (CHERRIES) to guide survey development and reporting of results.^17^ The survey (Supplementary file 1), designed by members of the research team, consisted of 61 questions including informed e-consent and occupational demographics (questions Q2-Q5) such as attributes of the speciality, provider type, and affiliations of respondents. It was constructed around five themes related to screening practice (Q6-Q37), treatment of pre-cancerous lesions and cancer (Q38-Q42), telemedicine (Q43-Q47), over- and under-screening in the pre-COVID-19 era (Q48-Q51), and resumption of in-person practice (Q52-Q61). The first two themes covered a range of questions that reflected the continuum of care in cervical cancer screening and management. The screening practice theme included sub-sections focusing on appointment scheduling (Q6-Q13), screening tests (Q14-Q21), human papillomavirus (HPV) self-sampling (Q22-Q23), colposcopy (Q24-Q29), and screening follow-up (Q30-Q37). Questions six through 47 were designed to collect data during the early COVID-19 period spanning from mid-March until mid-August 2020. For questions pertaining to the “resumption of in-person practice”, the period of interest was from mid-August 2020 until the date of survey completion. We also collected data on sex and age of the respondents. Respondents were asked to provide their impressions and best estimates when completing the survey, without necessarily confirming the proportions that they reported with their institution’s statistics.

For content validation and to determine question suitability and flow prior to the launch of the online survey, we conducted three iterative rounds of pilot testing by distributing the initial survey questionnaire to members of the Survey Study Group, consisting of 21 leading cervical cancer specialists and physicians in Canada who were not involved in study conception or design. Collective feedback in terms of relevance, appropriateness, and clarity of theme-related questions, as well as questionnaire length, was incorporated into the survey after each round; it was also used to refine the wording, type, and order of questions. Most were closed-ended (nominal, ordinal, and Likert-type questions), with few free-text questions and sub-questions that required elaborated responses. The study protocol and survey received ethical approval by the McGill Institutional Review Board on October 27, 2020.

### Survey administration and data management

The survey, constructed as a web-based questionnaire (originally developed in English and translated to French), was administered using LimeSurvey, an online-based survey tool hosted by McGill University. It was pretested by our research team and the panel of experts to ensure experiential functionality and valid data collection.

Several professional societies advertised and disseminated the survey via their email newsletter to respective members. These included the Society of Canadian Colposcopists (SCC; first email sent on November 18, 2020), Society of Gynecologic Oncology of Canada (GOC; first email sent on November 27, 2020), Canadian Association of Pathologists (CAP; first email sent on November 18, 2020), and Society of Obstetricians and Gynecologists of Canada (SOGC; first email sent on January 18, 2021). We reached out to primary care providers (i.e., family physicians, as well as general and nurse practitioners) via MDBriefCase (first email sent on February 7, 2021), which provides online continuing professional development to healthcare practitioners. Other platforms were also utilized to reach our targeted population, including posting a link to participate on the McGill Department of Family Medicine website, sending a request for participation letter containing the link to the listserv of the Chairs of Departments of Family Medicine across Canada, and social media platforms (LinkedIn and Twitter). The bilingual invitations to participate contained an email link to the web-based questionnaire survey. Two reminder emails – reiterating the objectives of the survey, inclusion criteria, and the survey link – were sent periodically (every 3-4 weeks).

Upon first entry to the survey portal, an informed e-consent form included a description of the survey, its objectives, and assurance of confidentiality of survey responses. Respondents who did not provide e-consent were unable to proceed to the survey questions. The platform allowed respondents to navigate between the different themes to revise their answers, if needed. Respondents received a $5 Starbucks gift card incentive upon completion of the survey, conditional on providing a valid email address.

The survey data, collected anonymously (no personal identifiers or IP addresses), were imported from LimeSurvey and curated in Excel (data cleaning and validation). We applied a two-step process to determine inclusion, the first based on eligibility criteria (target population) and the second on quality checks by flagging suspicious responses in open-ended questions. These questions entailed providing justification in Q22 and Q23 (if yes, no, or maybe, briefly justify); specification in Q47, Q54 (if other, specify) and Q61; description in Q60 (if yes, briefly describe); and any additional comments at the end of the survey.

Analyses included descriptive statistics and summaries of responses by province/territory (Alberta, British Columbia, Ontario, all other provinces, and territories), profession [primary care (i.e., general practitioners, family physicians, nurse practitioners and physician assistants); secondary care involving clinical diagnosis (i.e., colposcopists, and colposcopy registered nurses/register nurse practitioners); secondary care involving cytopathological diagnosis (i.e., cytopathologists and pathologists); and tertiary care involving gynecology and related activities (i.e., gynecological oncologists, gynecologists, obstetrician-gynecologists and gynecology nurses)], and place of practice (university-affiliated hospital, community hospital, public clinic, private clinic, community health center, and other). When responses to a given question were incomplete, we used the total number of complete responses as the denominator. Open-ended questions were analyzed using content analysis. Excel and SAS version 9.4 were used for data cleaning/visualization and analysis, respectively.

## Results

### Survey administration and responses

As shown in **Figure 1**, 778 potentially eligible respondents clicked on the survey link. Of those who started the survey, 235 were excluded as they were non-Canadian, had non-valid professions or places of practice, left the survey blank, or only completed the demographic section. Another 33 surveys were considered questionable; respondents took the survey multiple times, gave multiple non sequitur or contradictory answers, or plagiarized responses (copied text from websites/internet Google search). The final analysis sample comprised answers from 510 individuals, among whom the median time spent to complete the survey was 11 minutes and 53 seconds (interquartile range 6 minutes and 52 seconds – 17 minutes and 46 seconds). The survey was completed between 16 November 2020 and 28 February 2021.

**Figure 1:**
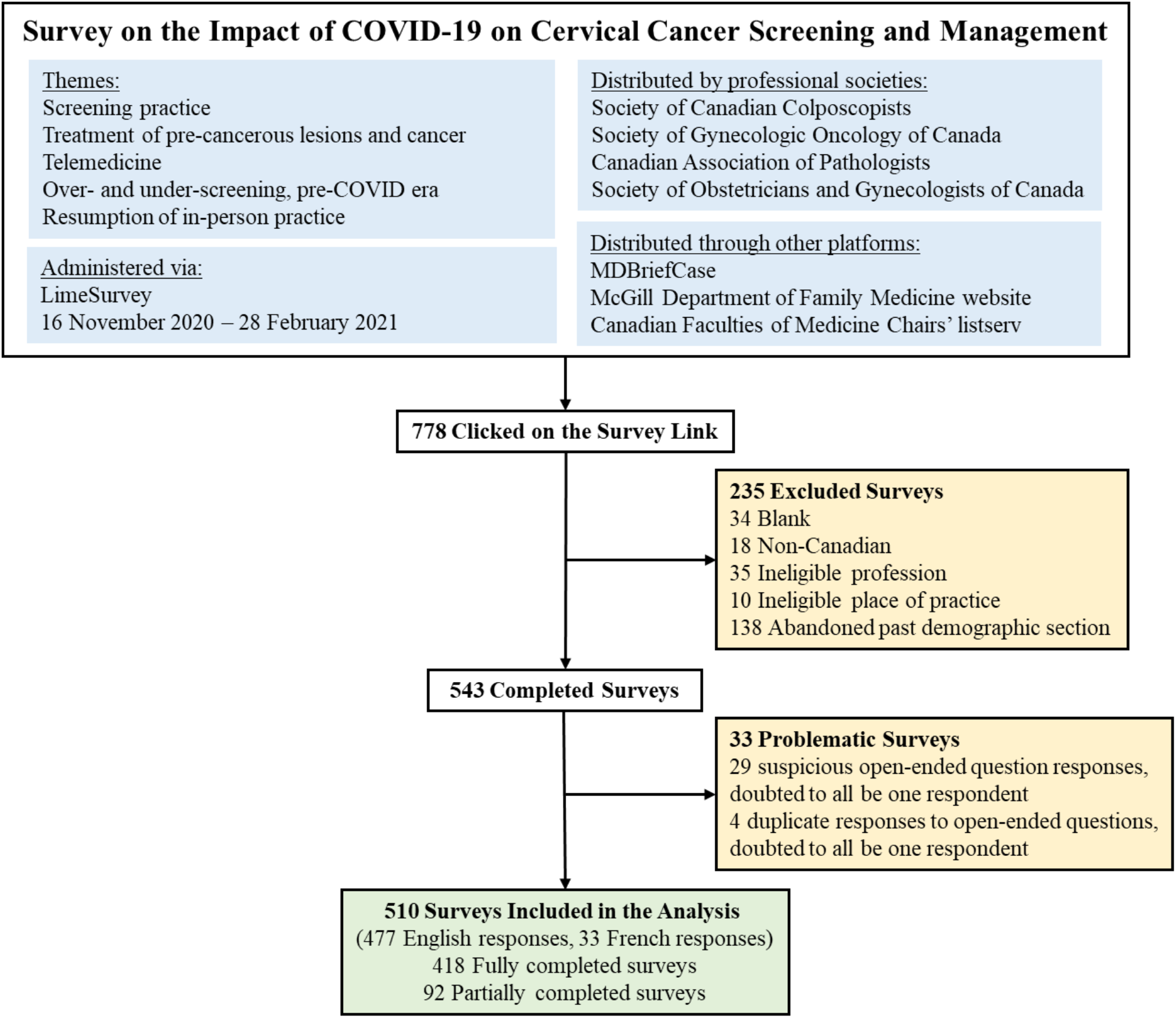
Description of survey elements and administration, and respondent flowchart

### Characteristics of survey respondents

**Table 1** summarizes the characteristics of the study population. There were more female than male respondents. The mean age was 44.4 years ± 11.9 (range 20-86, median: 42 years, interquartile range 35-54). Responses were mainly from Ontario, followed by British Columbia and Alberta. Most respondents were general practitioners/family physicians (43.7%), gynecologists/obstetrician-gynecologists (21.6%), nurse practitioners/registered nurses (14.1%), and colposcopists (10.2%). Regarding the place of practice, 32.9% reported working in private clinics, whereas comparable proportions reported working in university-affiliated hospitals (24.3%), community-affiliated hospitals (27.8%), and public clinics (25.3%). Some respondents selected multiple professions and/or places of practice. Of note, 42 of the 52 colposcopists were also gynecologists/obstetrician-gynecologists. Of the 124 respondents who practice in a university-affiliated hospital, 14, 11, and 16 respondents also selected community hospital, public clinic, and private clinic as a place of practice, respectively. Additionally, of the 142 respondents who work in a community hospital, 17 stated practicing in a public clinic and 23 in a private clinic.

**Table 1.** Characteristics of survey respondents (n = 510)

| **Variable** | **Categories** | **n (%)** |
| --- | --- | --- |
| **Sex** | Female | 284 (55.7) |
|  | Male | 124 (24.3) |
|  | *Not reported* | 102 (20.0) |
| **Age** | 20-29 | 26 (5.1) |
|  | 30-39 | 122 (23.9) |
|  | 40-49 | 98 (19.2) |
|  | 50-59 | 70 (13.7) |
|  | 60-69 | 42 (8.2) |
|  | 70+ | 7 (1.4) |
|  | *Not reported* | 145 (28.4) |
| **Province/Territory** | Alberta | 65 (12.8) |
|  | British Columbia | 107 (21.0) |
|  | Manitoba | 18 (3.5) |
|  | New Brunswick | 19 (3.7) |
|  | Newfoundland and Labrador | 7 (1.4) |
|  | Northwest Territories | 9 (1.8) |
|  | Nova Scotia | 21 (4.1) |
|  | Nunavut | 4 (0.8) |
|  | Ontario | 209 (41.0) |
|  | Prince Edward Island | 1 (0.2) |
|  | Quebec | 2 (0.4) |
|  | Saskatchewan | 21 (4.1) |
|  | Yukon | 26 (5.1) |
|  | *Not reported* | 1 (0.2) |
| **Profession\*** | Colposcopist | 52 (10.2) |
|  | Colposcopy Registered Nurse/Registered Practical Nurse | 16 (3.1) |
|  | Cytopathologist/technologist | 44 (8.6) |
|  | General Practitioner/Family Physician | 223 (43.7) |
|  | Gynecologist/Obstetrician-Gynecologist | 110 (21.6) |
|  | Gynecology Oncologist | 32 (6.3) |
|  | Gynecology Nurse | 21 (4.1) |
|  | Nurse Practitioner/Registered Nurse | 72 (14.1) |
|  | Pathologist | 17 (3.3) |
|  | Physician Assistant | 7 (1.4) |
|  | Other (manager in a community health center) | 1 (0.2) |
| **Place of Practice\*** | University-affiliated hospital | 124 (24.3) |
|  | Community hospital | 142 (27.8) |
|  | Public clinic | 129 (25.3) |
|  | Private clinic | 168 (32.9) |
|  | Community health center | 37 (7.3) |
|  | Other [homeless shelter (nurse); private lab (cytotechnologist)] | 2 (0.4) |

\*Frequency count exceeded number of respondents (510) as some selected more than 1 answer.

#### Theme 1: Screening practice

Cancellations and postponements of screening appointments were reported by 63.7% and 74.9% of respondents, respectively (**Table 2**). These are characterized in **Figure 2** by province (largely reported by healthcare professionals in Ontario), profession (largely reported by those in primary settings), and place of practice (largely reported by those in private clinics). Of the 325 respondents who reported cancellations of appointments, 55.7% stated that up to 49% of these appointments were cancelled by the physician or provider’s institution (**Table 2**). Similarly, 63.7% and 40.6% reported that up to 49% were cancelled by the patient or converted to telemedicine, respectively. Of the 382 healthcare professionals who reported that appointments were postponed, 51.6%, 68.4%, and 42.9% respectively stated that up to 49% of these appointments were postponed by the physician or provider’s institution, by the patient, or converted to telemedicine. The majority of appointments (64.4%) were at most deferred by less than 4 months, whereas 9.4% were deferred by more than 6 months. **Supplementary Figures 1-3** show the proportions of all cancelled and postponed screening appointments by province, profession, and place of practice, respectively. Likewise, most responses were from professionals in Ontario, primary care, and private clinics. A total of 99 respondents (19.4%) reported that their practice/institution did not allow in person consultation appointments during the pandemic’s peak period (**Supplementary Table 1**). Of those who reported allowance of in person consultations (378, 74.1%), most were from Ontario, in primary care, and practicing in private clinics (**Figure 4**).

**Table 2.**
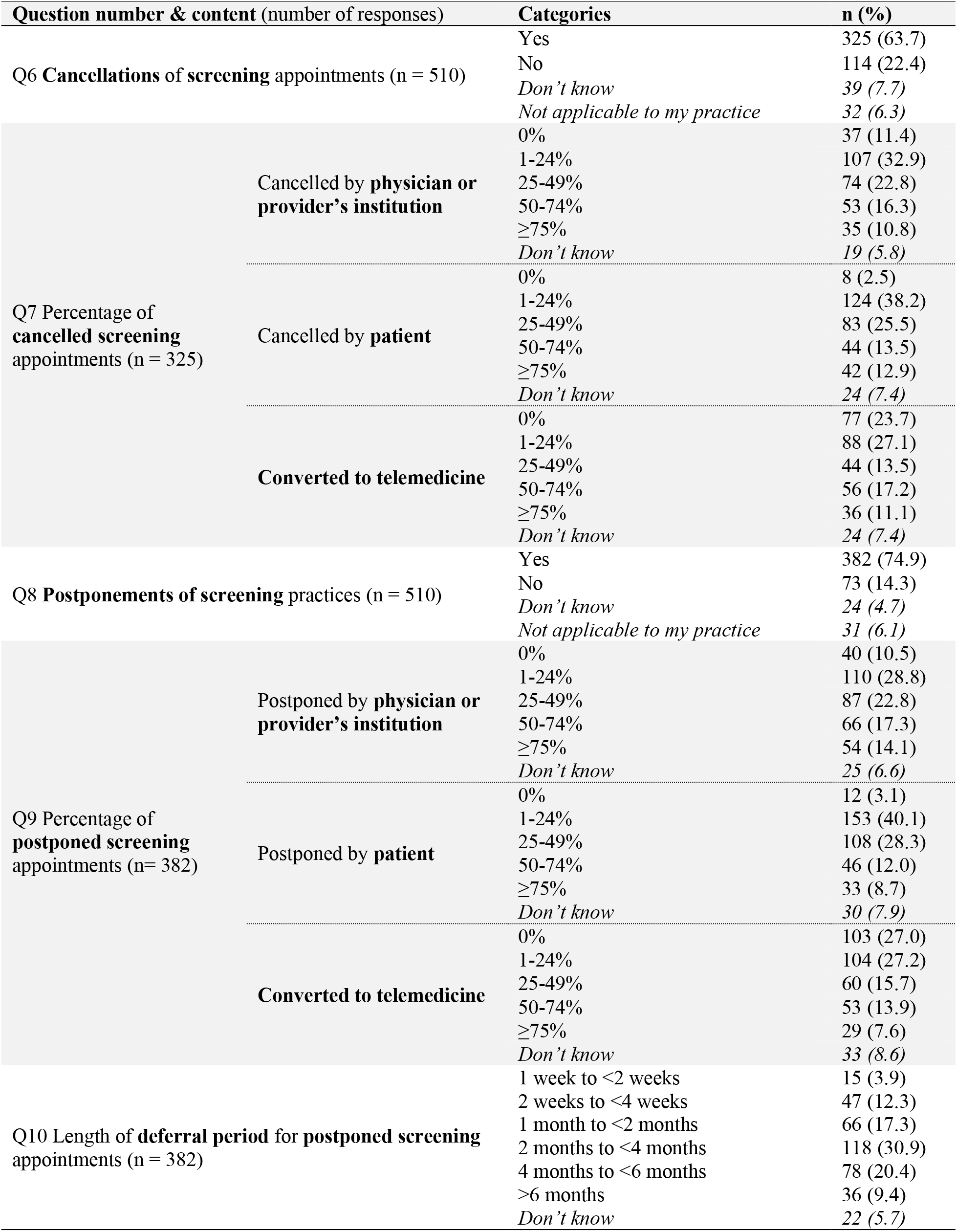
Cancellations and postponements of cervical cancer screening appointments

**Figure 2:**
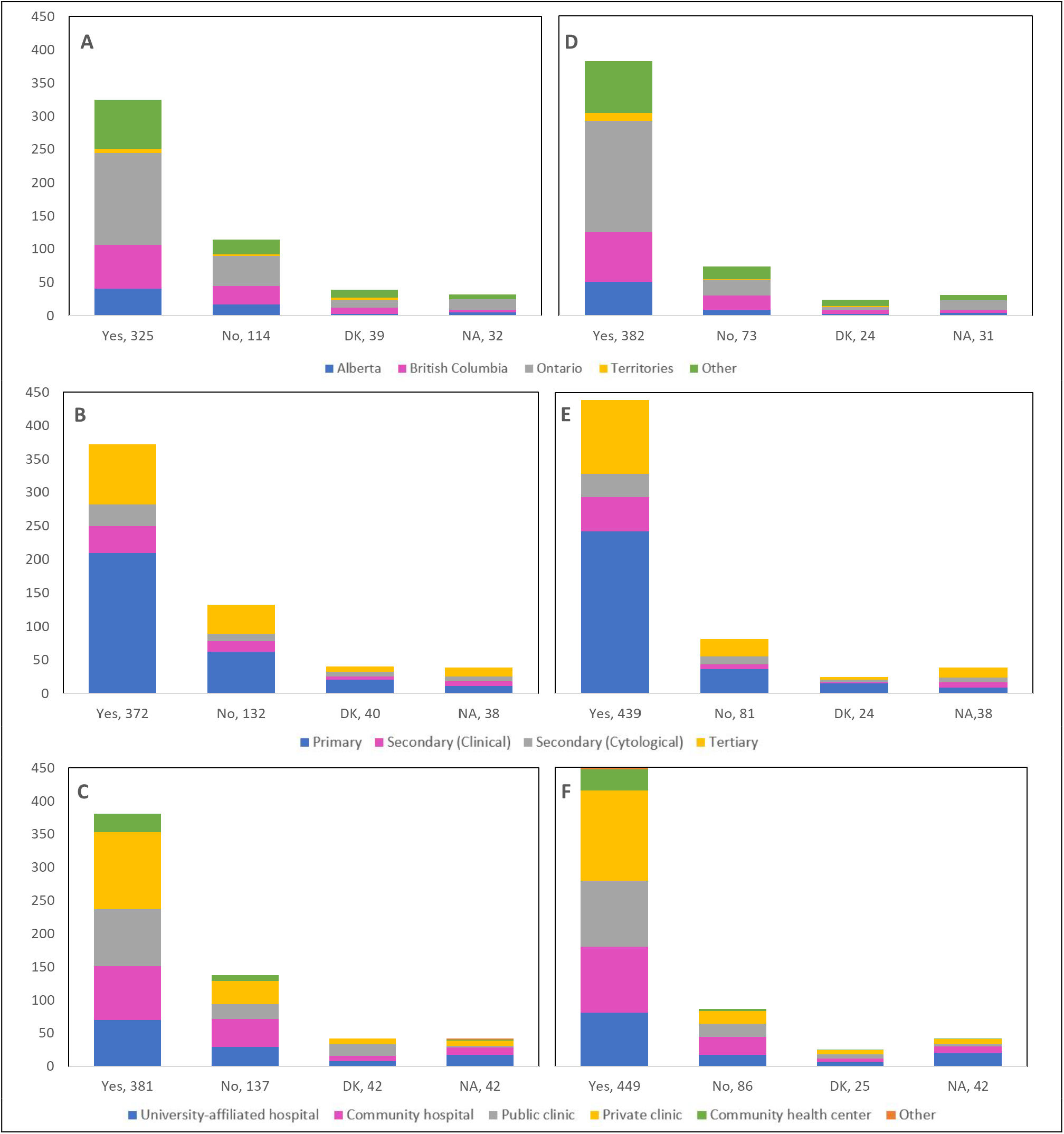
Cancellations and postponements of **screening** appointments by province, profession, and place of practice (n = 510) Number of <u>cancellations</u> are shown by (A) province, (B) profession, and (C) place of practice. Number of <u>postponements</u> are shown by (D) province, (E) profession, and (F) place of practice. Answers include responses for questions 6 (cancellations) and 8 (postponements) by questions 2 (province), 4 (profession), and 5 (place of practice). **Panels A and D:** Territories include Northwest Territories, Nunavut, and Yukon. Other provinces include Manitoba, New Brunswick, Newfoundland and Labrador, Nova Scotia, Prince Edward Island, Quebec, and Saskatchewan (and one respondent who preferred not to say). **Panels B and E:** Primary includes general practitioners/family physicians, nurse practitioners/registered nurses, physician assistants and a manager of a community health center; Secondary (clinical) includes colposcopists and colposcopy registered nurses/registered practical nurses; Secondary (cytological) includes cytopathologists/technologists and pathologists; Tertiary includes gynecologists/obstetrician-gynecologists, gynecology oncologists, and gynecology nurses. **Panels B, C, E and F:** Frequency count exceeded total number of respondents as some reported multiple professions and places of practice. DK: Don’t know; NA: Not applicable to my practice

**Figure 3:**
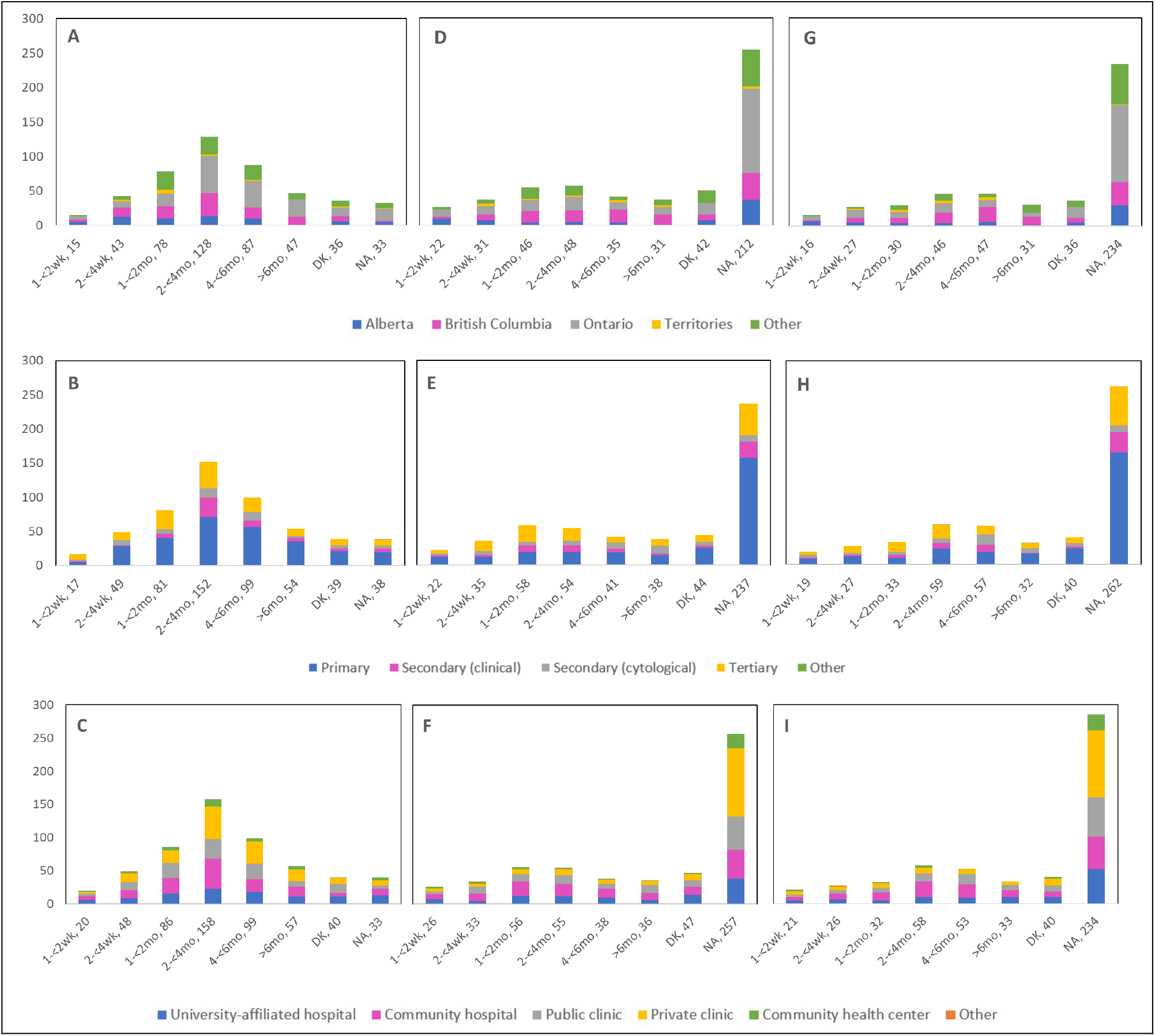
Deferral period for **postponed screening** appointments by province, profession, and place of practice (n = 467) Deferral period for postponed <u>Pap test</u> appointments is shown by (A) province, (B) profession, and (C) place of practice. Deferral period for postponed <u>HPV test</u> appointments is shown by (D) province, (E) profession, and (F) place of practice. Deferral period for postponed <u>HPV/Pap co-test</u> appointments is shown (G) province, (H) profession, and (I) place of practice. Answers include responses for question 19 by questions 2 (province), 4 (profession) and 5 (place of practice). **Panels A, D and G:** Territories include Northwest Territories, Nunavut, and Yukon. Other provinces include Manitoba, New Brunswick, Newfoundland and Labrador, Nova Scotia, Prince Edward Island, Quebec, and Saskatchewan (and one respondent who preferred not to say). **Panels B, E and H:** Primary includes general practitioners/family physicians, nurse practitioners/registered nurses, physician assistants, and a manager of a community health center; Secondary (clinical) includes colposcopists and colposcopy registered nurses/registered practical nurses; Secondary (cytological) includes cytopathologists/technologists and pathologists; Tertiary includes gynecologists/obstetrician-gynecologists, gynecology oncologists, and gynecology nurses. **Panels B, C, E, F, H and I:** Frequency count exceeded total number of respondents as some reported multiple professions and places of practice. DK: Don’t know; NA: Not applicable to my practice

**Figure 4:**
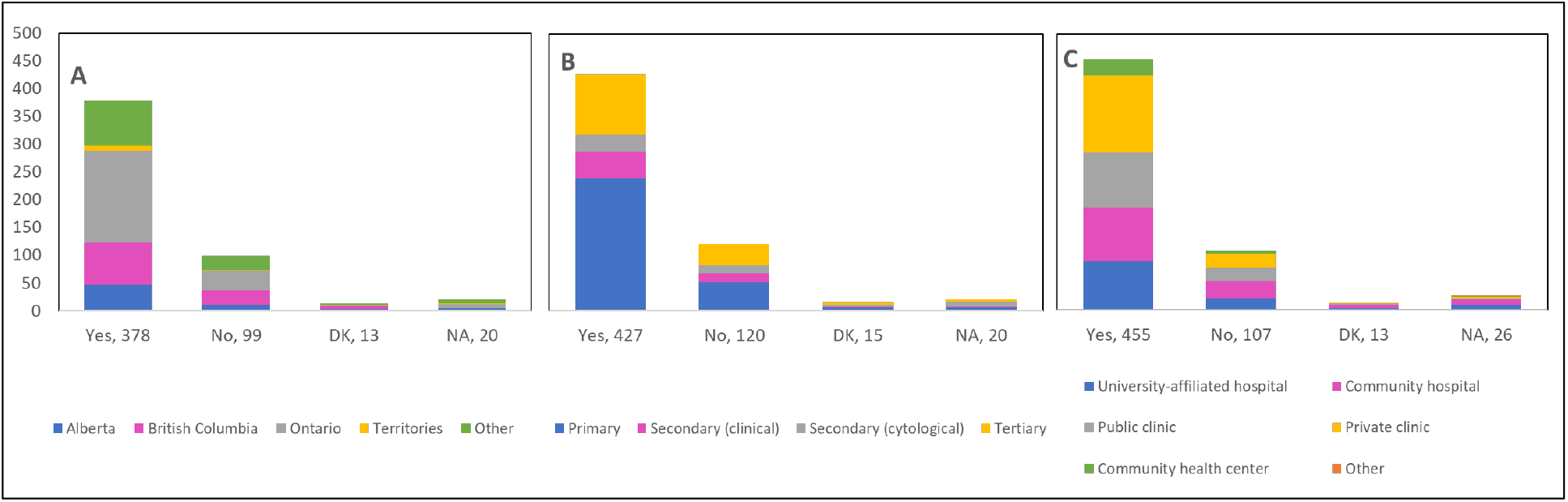
Allowance of **in person consultations** during the peak period of the pandemic by province, profession, and place of practice (n = 510) Number of <u>in person consultations</u> is shown by (A) province, (B) profession, and (C) place of practice. Answers include responses for question 11 by questions 2 (province), 4 (profession) and 5 (place of practice). **Panel A:** Territories include Northwest Territories, Nunavut, and Yukon. Other provinces include Manitoba, New Brunswick, Newfoundland and Labrador, Nova Scotia, Prince Edward Island, Quebec, and Saskatchewan (and one respondent who preferred not to say). **Panel B:** Primary includes general practitioners/family physicians, nurse practitioners/registered nurses, physician assistants and a manager of a community health center; Secondary (clinical) includes colposcopists and colposcopy registered nurses/registered practical nurses; Secondary (cytological) includes cytopathologists/technologists and pathologists; Tertiary includes gynecologists/obstetrician-gynecologists, gynecology oncologists, and gynecology nurses. **Panels B and C:** Frequency count exceeded total number of respondents as some reported multiple professions and places of practice. DK: Don’t know; NA: Not applicable to my practice

In terms of the type of test usually employed for primary cervical cancer screening (**Supplementary Table 2**), 76.1% of respondents reported cytology, 32.8% the HPV test, and 25.4% reported using both. Compared to pre-COVID-19, 15.8%, 4.9%, and 3.8% reported a decrease by 75% or more in the number of Pap, HPV, and co-tests, respectively. Delays in scheduling of these tests were correspondingly reported by 56.9%, 22.6%, and 21.8% of respondents. Of the 469 healthcare professionals who reported cancellations of a scheduled screening test, 48.1%, 19.8%, and 17.3% stated that up to 49% of Pap, HPV, and co-tests were cancelled, whereas of the 468 professionals who reported postponements, the corresponding proportions were 46.8%, 22.3%, and 20.1% (**Supplementary Table 3**). Pap tests (56.5%), HPV tests (31.5%) and HPV/Pap co-tests (25.5%) were deferred by less than 4 months, at the most, whereas 10.1%, 6.6%, and 6.6% of these tests were deferred by more than 6 months, respectively. **Figure 3** illustrates the deferral period of these postponed screening tests appointments by province, profession, and place of practice. Regarding the delay in forwarding tests to the laboratory, 15%, 15.6%, and 13.9% of respondents reported such delays for Pap, HPV, and co-tests, respectively (**Supplementary Table 4**).

When asked about whether the pandemic will encourage/facilitate/accelerate the implementation of HPV self-sampling in cervical cancer screening programs at a provincial and/or national level, 150/455 respondents (33%) indicated that it will, and 50.1% were in favor of implementing this modality as an alternative screening method (**Supplementary Table 5**).

With respect to colposcopy appointments, cancellations and postponements were reported by 25.2% and 37% of respondents (**Supplementary Table 6**), with patterns and changes by province, profession, and place of practice (**Figure 5, Supplementary Figures 4-6**) similar to those reported for screening appointments. The same was observed for reported cancellations (33.3%) and postponements (53.5%) of follow-up appointments (**Supplementary Table 7, Figure 6**, and **Supplementary Figures 7-9**). With respect to receiving test results from the lab prior to follow-up with patients, 21.1%, 19.3%, and 12.1% of respondents reported delays for Pap, HPV, and co-tests, respectively (**Supplementary Table 8**).

**Figure 5:**
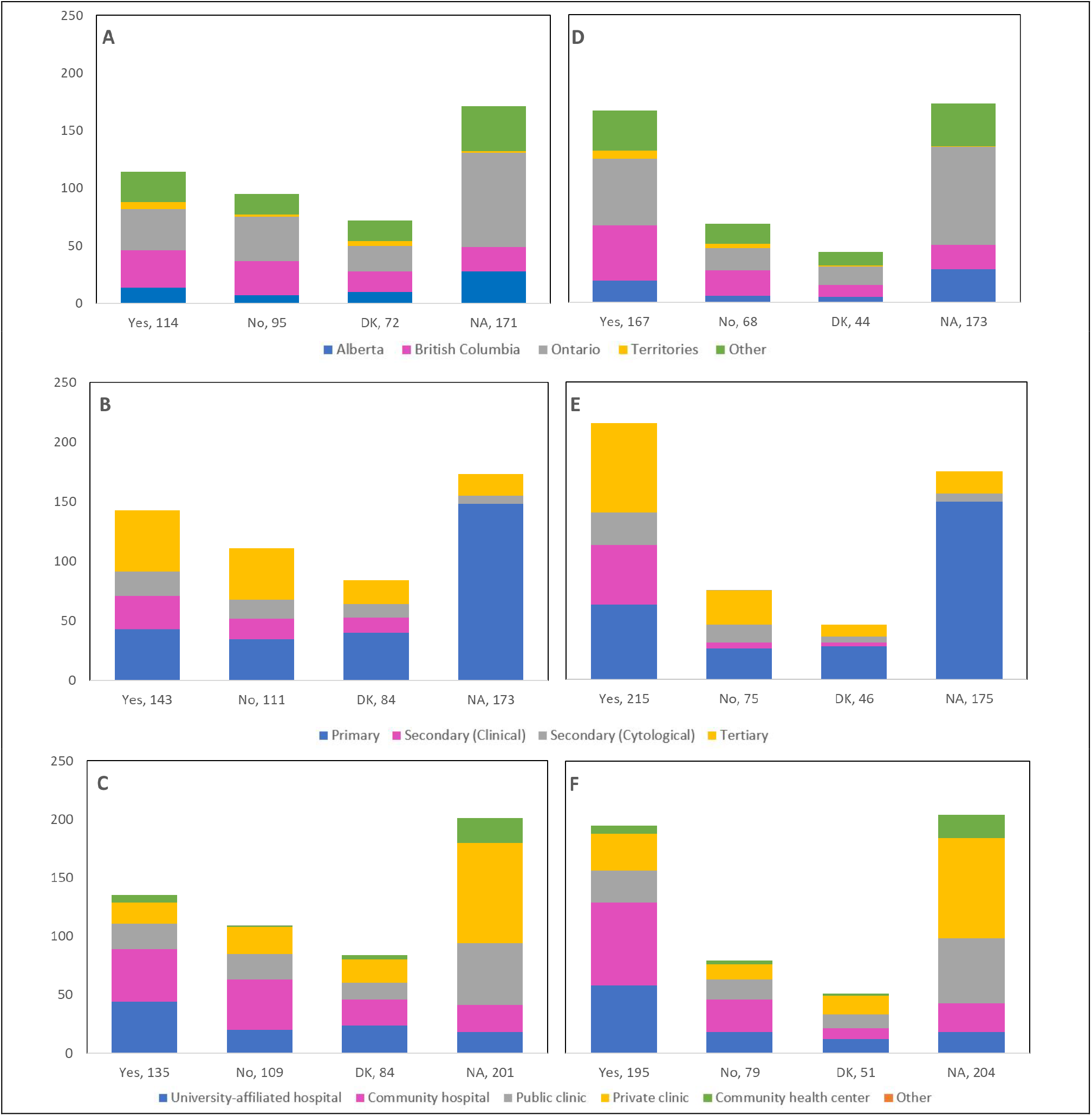
Cancellations and postponements of **colposcopy** appointments by province, profession, and place of practice (n = 452) Number of <u>cancellations</u> are shown by (A) province, (B) profession, and (C) place of practice. Number of <u>postponements</u> are shown by (D) province, (E) profession, and (F) place of practice. Answers include responses for questions 25 (cancellations) and 27 (postponements) by questions 2 (province), 4 (profession), and 5 (place of practice). **Panels A and D:** Territories include Northwest Territories, Nunavut, and Yukon. Other provinces include Manitoba, New Brunswick, Newfoundland and Labrador, Nova Scotia, Prince Edward Island, Quebec, and Saskatchewan (and one respondent who preferred not to say). **Panels B and E:** Primary includes general practitioners/family physicians, nurse practitioners/registered nurses, physician assistants, and a manager of a community health center; Secondary (clinical) includes colposcopists and colposcopy registered nurses/registered practical nurses; Secondary (cytological) includes cytopathologists/technologists and pathologists; Tertiary includes gynecologists/obstetrician-gynecologists, gynecology oncologists, and gynecology nurses. **Panels B, C, E and F:** Frequency count exceeded total number of respondents as some reported multiple professions and places of practice. DK: Don’t know; NA: Not applicable to my practice

**Figure 6:**
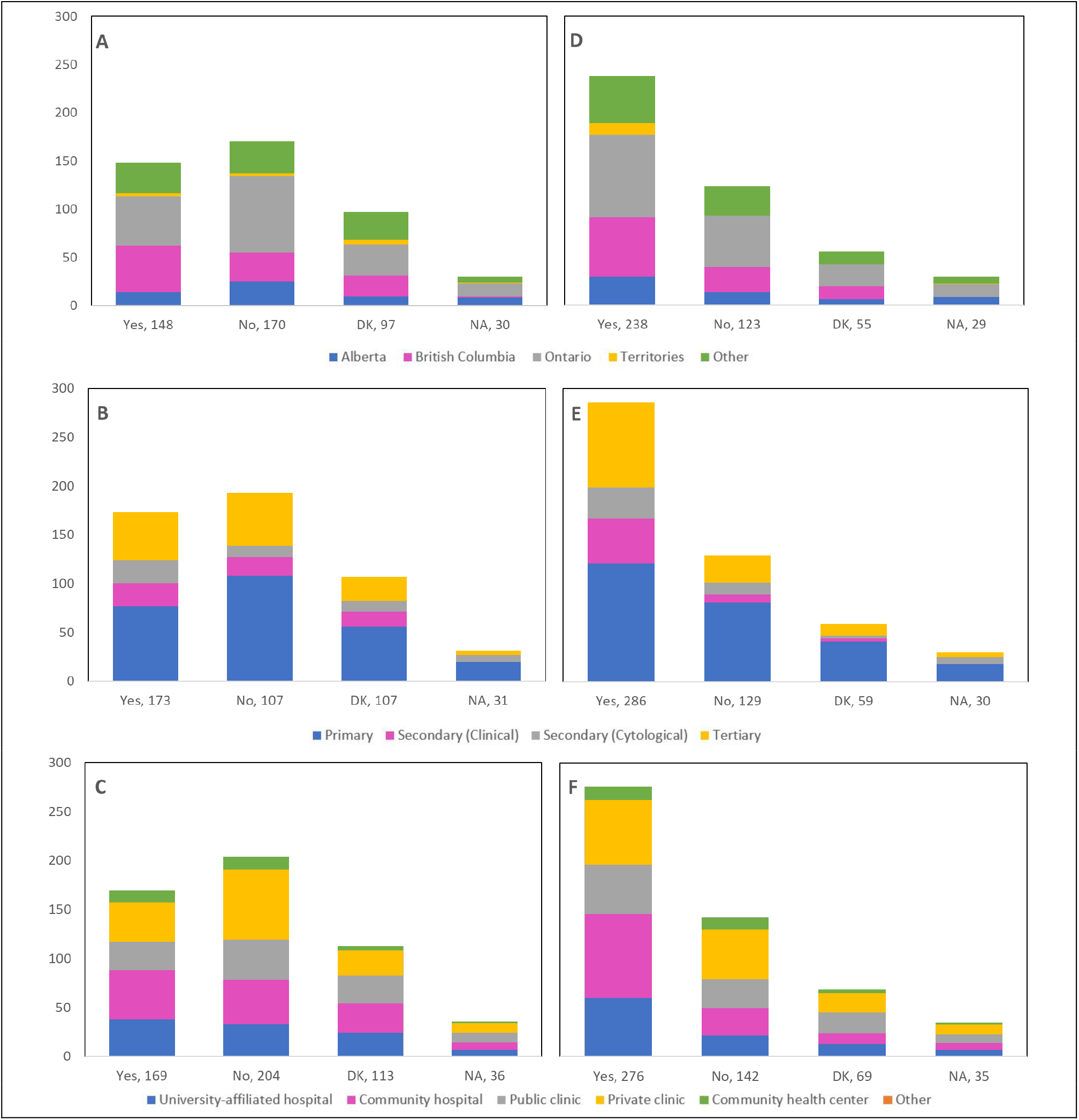
Cancellations and postponements of **follow-up** appointments by province, profession, and place of practice (n = 445) Number of <u>cancellations</u> are shown by (A) province, (B) profession, and (C) place of practice. Number of <u>postponements</u> are shown by (D) province, (E) profession, and (F) place of practice. Answers include responses for questions 31 (cancellations) and 33 (postponements) by questions 2 (province), 4 (profession), and 5 (place of practice). **Panels A and D:** Territories include Northwest Territories, Nunavut, and Yukon. Other provinces include Manitoba, New Brunswick, Newfoundland and Labrador, Nova Scotia, Prince Edward Island, Quebec, and Saskatchewan (and one respondent who preferred not to say). **Panels B and E:** Primary includes general practitioners/family physicians, nurse practitioners/registered nurses, physician assistants, and a manager of a community health center; Secondary (clinical) includes colposcopists and colposcopy registered nurses/registered practical nurses; Secondary (cytological) includes cytopathologists/technologists and pathologists; Tertiary includes gynecologists/obstetrician-gynecologists, gynecology oncologists, and gynecology nurses. **Panels B, C, E and F:** Frequency count exceeded total number of respondents as some reported multiple professions and places of practice. DK: Don’t know; NA: Not applicable to my practice

#### Theme 2: Treatment of pre-cancerous lesions and cancer

**Supplementary Table 9** presents the observed changes in the number of treatment procedures by treatment type reported by 431 respondents; cold knife conization (15.8% decrease, 12.5% unaffected, 15.8% increase), other excisional (20.0% decrease, 12.8% unaffected, 15.5% increase), ablative procedures (15.8% decrease, 10.9% unaffected, 16.3% increase), hysterectomy (23.9% decrease, 9.3% unaffected, 13.4% increase), chemotherapy (10.9% decrease, 10.9% unaffected, 15.1% increase), and radiation (13.7% decrease, 7.9% unaffected, 14.0% increase). The number of cancellations or postponements of treatment procedures (13.5% for cold knife conization, 23.7% for other excisional, 19.5% for ablative procedures, 21.1% for hysterectomy, 10.4% for chemotherapy, and 11.6% for radiation) are shown by province (**Figure 7**), profession (**Figure 8**), and place of practice (**Figure 9**).

**Figure 7:**
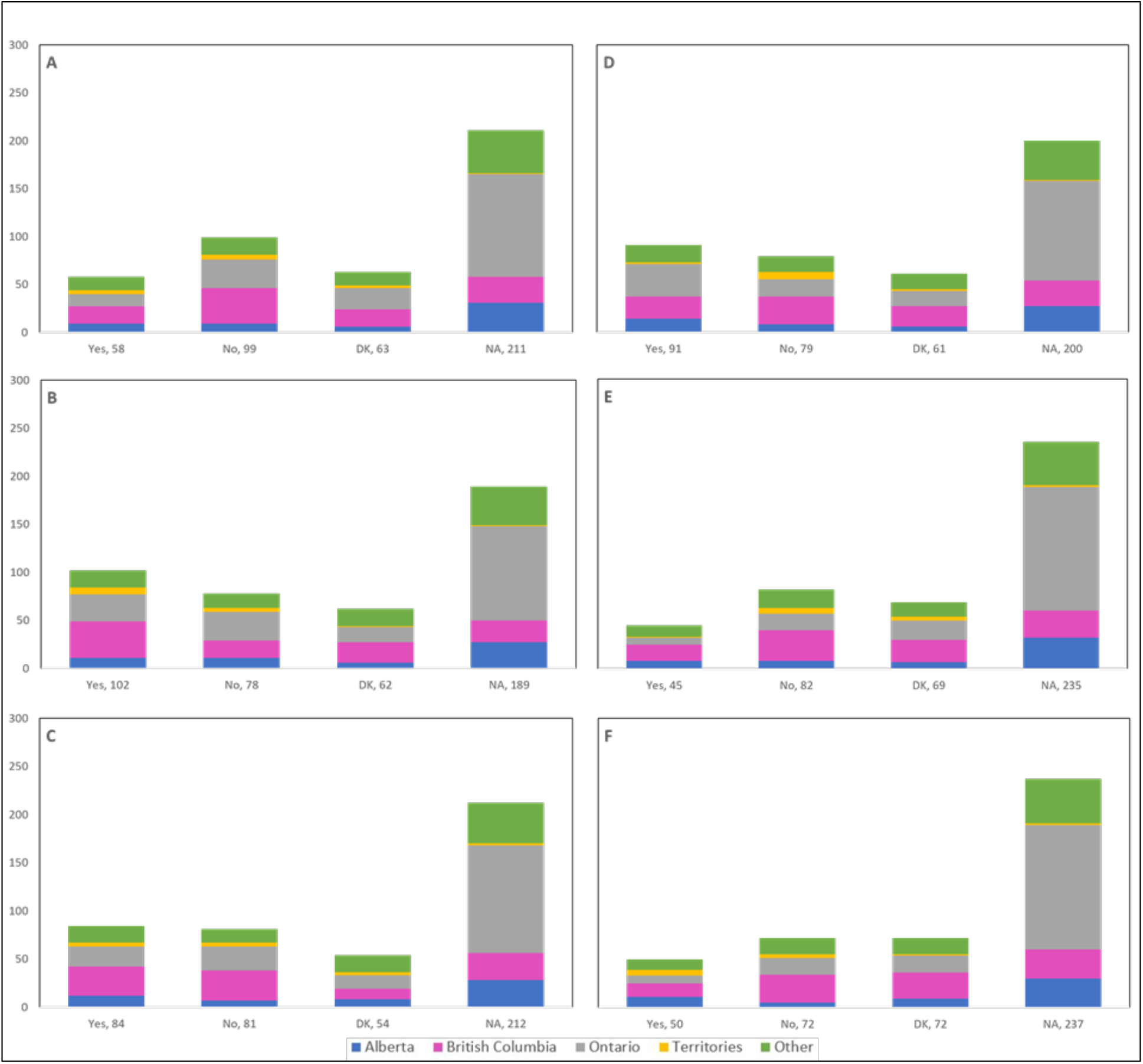
Cancellations or postponements of **treatment procedures** by **province** (n = 431) Number of <u>cancellations or postponements</u> of (A) Cold Knife Conization, (B) Other excisional (e.g., LEEP), (C) Ablative procedures, (D) Hysterectomy, (E) Chemotherapy, and (F) Radiation are shown by province. Answers include the responses for question 39 by question 2. Territories include Northwest Territories, Nunavut, and Yukon. Other provinces include Manitoba, New Brunswick, Newfoundland and Labrador, Nova Scotia, Prince Edward Island, Quebec, and Saskatchewan (and one respondent who preferred not to say). DK: Don’t know; NA: Not applicable to my practice

**Figure 8:**
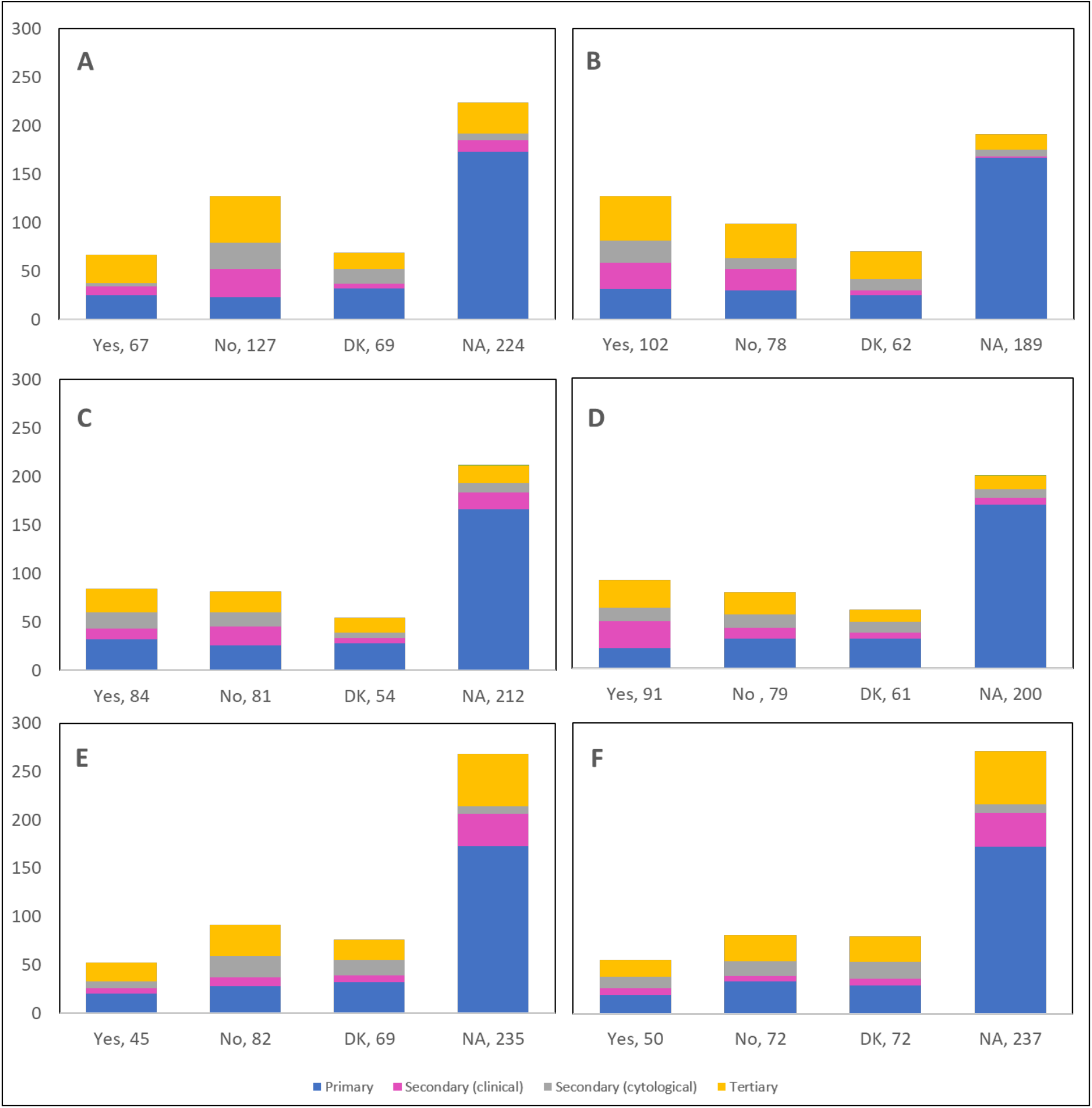
Cancellations or postponements of **treatment procedures** by **profession** (n = 431) Number of <u>cancellations or postponements</u> of (A) Cold Knife Conization, (B) Other excisional (e.g., LEEP), (C) Ablative procedures, (D) Hysterectomy, (E) Chemotherapy, and (F) Radiation are shown by profession. Answers include the responses for question 39 by question 4. Primary includes general practitioners/family physicians, nurse practitioners/registered nurses, physician assistants, and a manager of a community health center; Secondary (clinical) includes colposcopists and colposcopy registered nurses/registered practical nurses; Secondary (cytological) includes cytopathologists/technologists and pathologists; Tertiary includes gynecologists/obstetrician-gynecologists, gynecology oncologists, and gynecology nurses. Frequency count exceeded total number of respondents as some reported multiple professions. DK: Don’t know; NA: Not applicable to my practice

**Figure 9:**
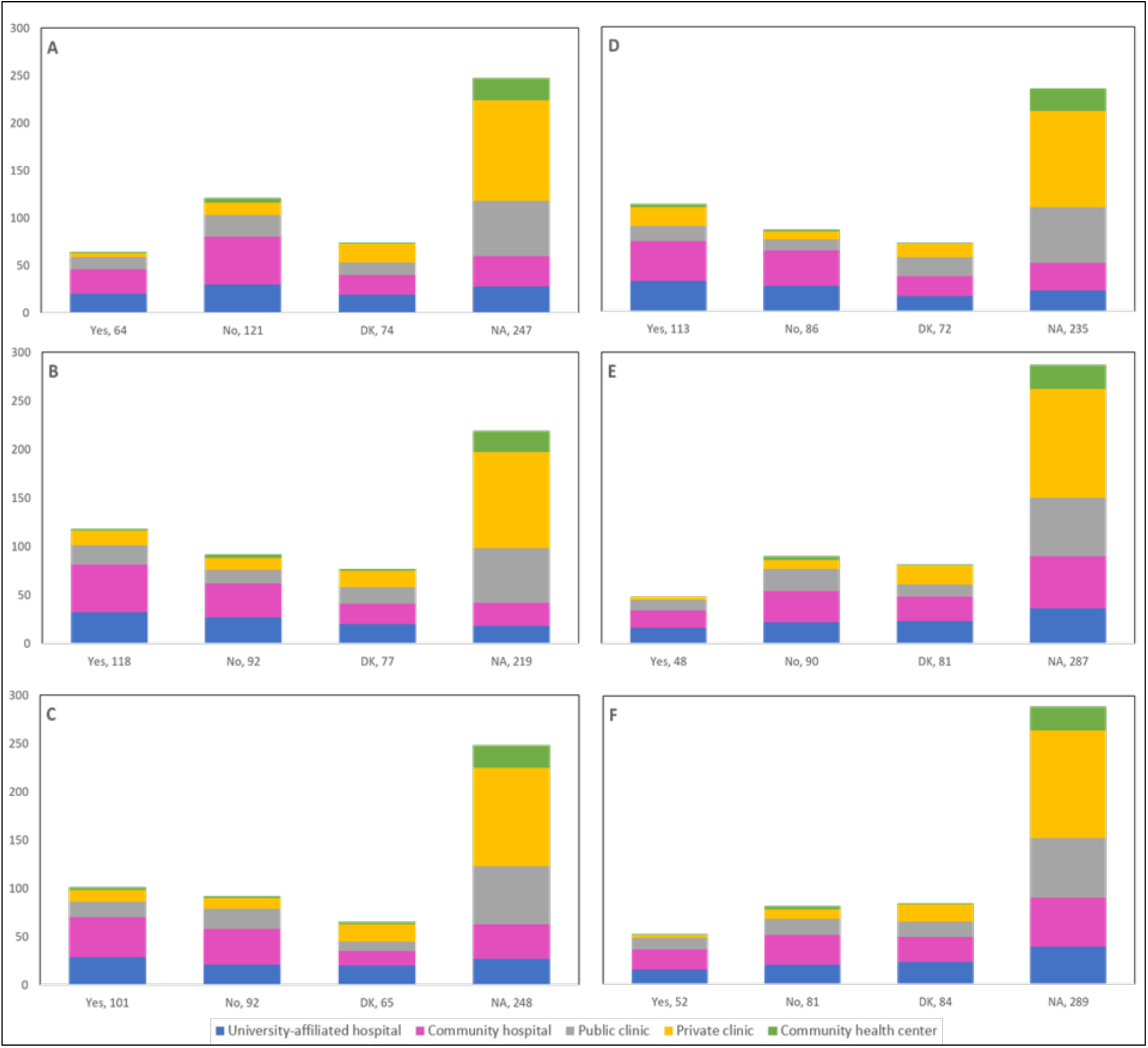
Cancellations or postponements of **treatment procedures** by **place of practice** (n = 431) Number of <u>cancellations or postponements</u> of (A) Cold Knife Conization, (B) Other excisional (e.g., LEEP), (C) Ablative procedures, (D) Hysterectomy, (E) Chemotherapy, and (F) Radiation are shown by place of practice. Answers include the responses for question 39 by question 5. Frequency count exceeded total number of respondents as some reported multiple places of practice. DK: Don’t know; NA: Not applicable to my practice

#### Theme 3: Telemedicine

**Table 3** presents responses reported by 429 respondents regarding the adoption of telemedicine. A total of 384 respondents (89.5%) reported that their practice/institution adopted telemedicine to communicate with patients; 26.8% indicated that they called 25-49% of their patients for distance consultations and 19.8% indicated the use of telemedicine with 25-49% of patients for follow-up appointments related to a cervical cancer screening procedure. Around two-thirds (72.7%) of healthcare professionals reported that virtual consultations are covered by their jurisdictional public health insurance system. Regarding which interactions with patients would be appropriate to convert to telemedicine, 82.1% of respondents selected test results reporting, 66% health and medical history reporting, 51.7% consent forms prior to in person procedures, 42.9% post-procedure follow-up, and 33.1% selected in person appointment planning/scheduling.

**Table 3.**
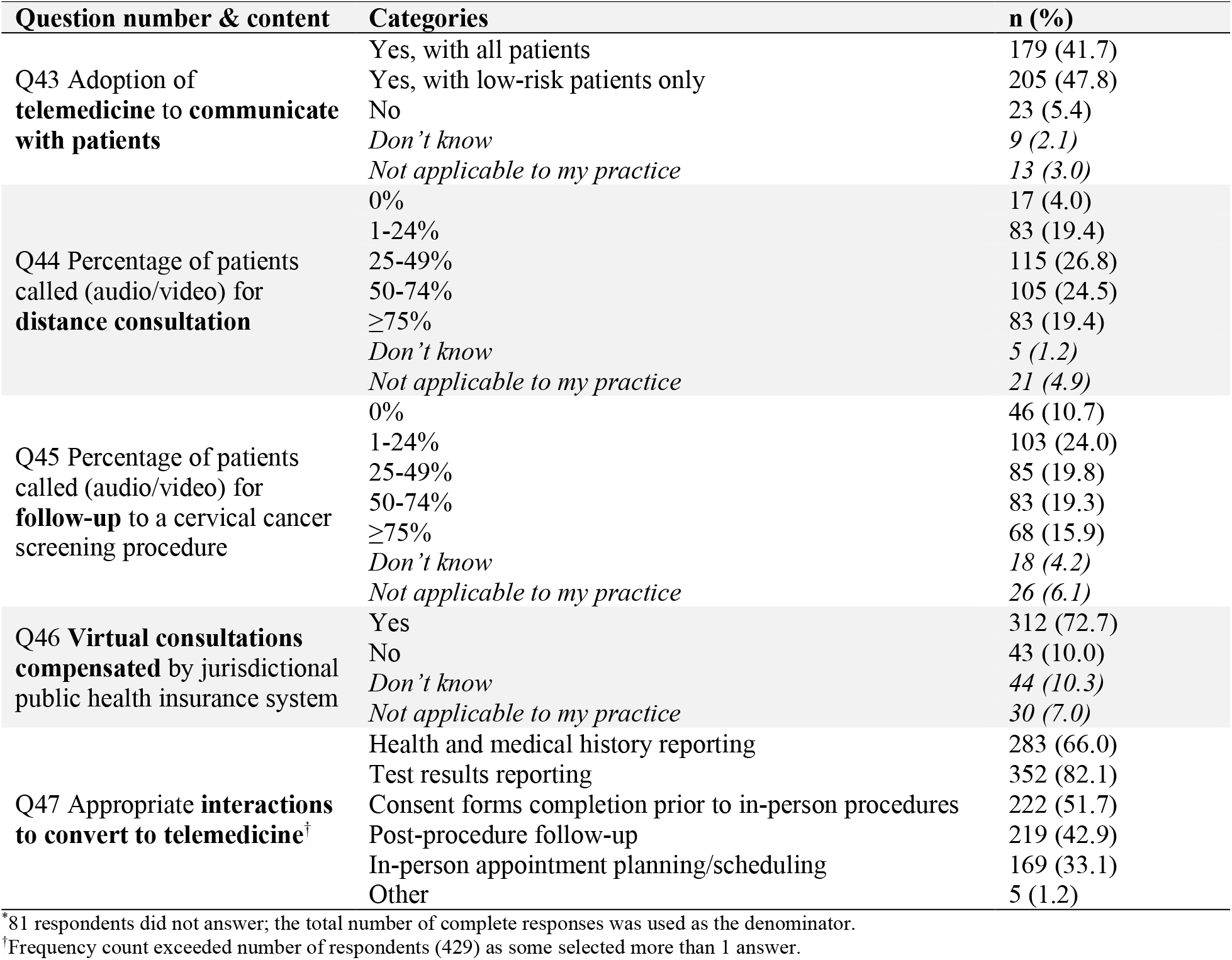
Adoption of telemedicine (n = 429^*^)

#### Theme 4: Over- and under-screening in the pre-COVID-19 era

There was a total of 190 responses (44.5%) indicating issues of over-screening/over-diagnosis/over-treatment prior to the onset of the pandemic, with over-diagnosis (20.1%) of cervical lesions being the most commonly reported issue (**Table 4**). A minority of respondents reported that the current delays/cancellations of screening and management procedures may have had a positive impact in reducing unnecessary screening (15.2% of responses), diagnosis (20.6% of responses), and treatment (10.5% of responses). Conversely, 350 responses (81.9%) indicated issues of under-screening/under-diagnosis/under-treatment pre-COVID-19, and in turn, reported that the current delays/cancellations of screening and management procedures may have had a negative impact by reducing necessary screening (47.8% of responses), diagnosis (48.4% of responses), and treatment (25.3% of responses).

**Table 4.**
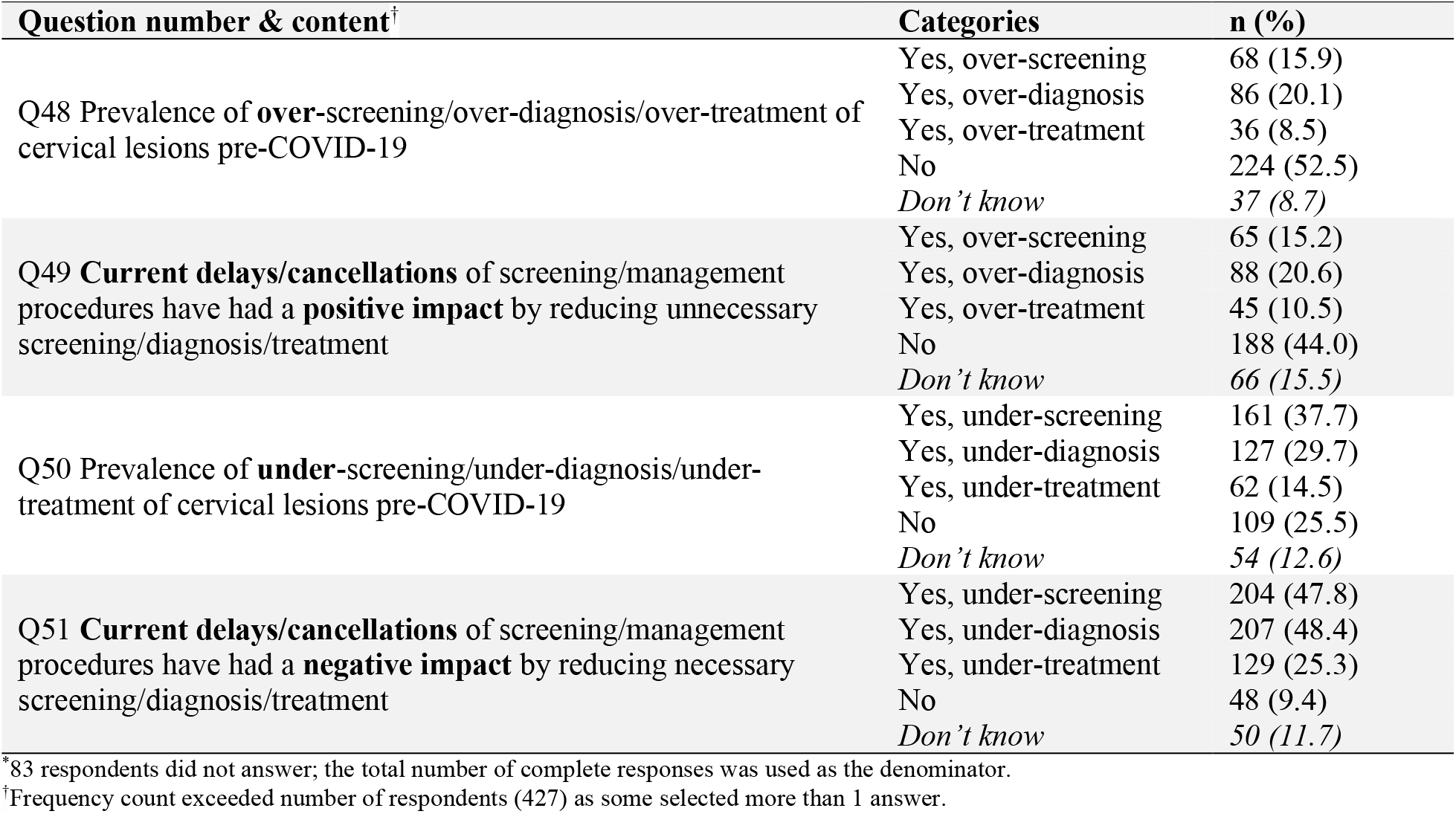
Over-screening and under-screening in the pre-COVID-19 era (n = 427^*^)

#### Theme 5: Resumption of in-person practice

Nearly half (45.1%) of respondents reported that their practice/institution has caught up with the cancellations/postponements of appointments caused by restrictions introduced at the beginning of the pandemic, whereas 34.9% reported ongoing disruptions and delays (**Table 5**). Allowing longer workdays and/or working on weekends, increasing availability of operating rooms for treatment procedures, and converting operating room procedures to take place in clinics constituted the main measures that were implemented to catch up with these cancellations/postponements. A total of 160 respondents (38%) indicated that their practice/institution has currently caught up with 50% or more of the cancellations/postponements. Nonetheless, 51.3% reported that patients have not been coming in for routine screening procedures at a capacity equivalent to the pre-COVID-19 era. Almost a third of respondents (29.2%) answered that 25-49% of patients have been attending routine screening procedures, in comparison to the pre-COVID-19 era. Most of these respondents were from Ontario, practicing in primary care settings at community hospitals (**Figure 10**). Notably, 32.3% mentioned an increase in the frequency of patients presenting with worsened symptoms, and 16.6% reported that 25-49% of patients had been diagnosed with more advanced cytological abnormalities and/or lesions confirmed by histology, compared to the pre-COVID-19 era **(Table 5)**.

**Table 5.** Resumption of in-person practice (n = 421^*^)

| Question number & content | Categories | n (%) |
| --- | --- | --- |
| Q53 Practice/institution **caught up with cancellations/postponements** | Yes | 190 (45.1) |
|  | No | 147 (34.9) |
|  | *Don't know* | 58 (13.8) |
|  | *Not applicable to my practice* | 26 (6.2) |
| Q54 Measures implemented to **catch up** with cancellations/postponements<sup>†</sup> | Allow longer workdays and/or working on weekends | 91 (21.6) |
|  | Increase availability of OR for treatment procedures | 91 (21.6) |
|  | Convert OR procedures, if possible, to take place in clinics | 74 (17.6) |
|  | Increase availability to labs for processing test samples | 48 (11.4) |
|  | Other | 51 (11.9) |
|  | *Don't know* | 41 (9.7) |
|  | *Not applicable to my practice* | 92 (21.9) |
| Q55 Percentage of **cancellations/postponements** currently **caught up with** | 0% | 5 (1.2) |
|  | 1-24% | 60 (14.3) |
|  | 25-49% | 98 (23.3) |
|  | 50-74% | 72 (17.1) |
|  | ≥75% | 88 (20.9) |
|  | *Don't know* | 55 (13.1) |
|  | *Not applicable to my practice* | 43 (10.2) |
| Q56 Patients **attending routine screening at equivalent capacity** to pre-COVID-19 era | Yes | 132 (31.4) |
|  | No | 216 (51.3) |
|  | *Don't know* | 52 (12.4) |
|  | *Not applicable to my practice* | 21 (5.0) |
| Q57 Percentage of patients attending **routine screening** compared to **pre-COVID-19** | 0% | 3 (0.7) |
|  | 1-24% | 75 (17.8) |
|  | 25-49% | 123 (29.2) |
|  | 50-74% | 89 (21.1) |
|  | ≥75% | 76 (18.1) |
|  | *Don't know* | 35 (8.3) |
|  | *Not applicable to my practice* | 20 (4.8) |
| Q58 **Increase in frequency** of patients with **worsening of symptoms** during screening | Yes | 136 (32.3) |
|  | No | 216 (51.3) |
|  | *Don't know* | 47 (11.2) |
|  | *Not applicable to my practice* | 22 (5.2) |
| Q59 Percentage of patients diagnosed with **more advanced cytological abnormalities/lesions**, in comparison to pre-COVID-19 | 0% | 98 (23.3) |
|  | 1-24% | 94 (22.3) |
|  | 25-49% | 70 (16.6) |
|  | 50-74% | 42 (10.0) |
|  | ≥75% | 6 (1.4) |
|  | *Don't know* | 95 (22.6) |
|  | *Not applicable to my practice* | 16 (3.8) |
| Q60 **Screening patients (with COVID-19)** for cervical cancer | Yes | 113 (26.8) |
|  | No | 222 (52.7) |
|  | *Don't know* | 58 (13.8) |
|  | *Not applicable to my practice* | 28 (6.7) |

\*89 respondents did not answer; the total number of complete responses was used as the denominator.
<sup>†</sup>Frequency count exceeded number of respondents (421) as some selected more than 1 answer.

**Figure 10:**
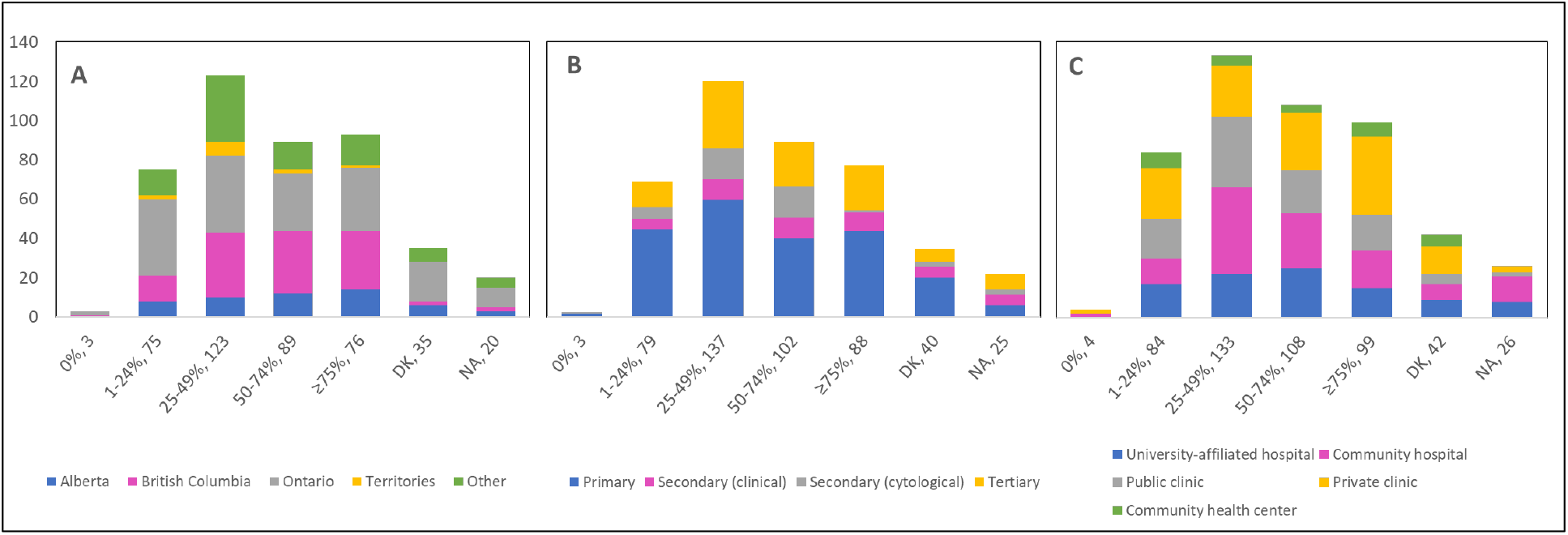
Percentage of patients attending routine screening compared to pre-COVID-19 by province, profession, and place of practice (n = 421) Proportions are shown by (A) province, (B) profession, and (C) place of practice. Answers include responses for question 57 by questions 2 (province), 4 (profession) and 5 (place of practice). **Panel A:** Territories include Northwest Territories, Nunavut, and Yukon. Other provinces include Manitoba, New Brunswick, Newfoundland and Labrador, Nova Scotia, Prince Edward Island, Quebec, and Saskatchewan (and one respondent who preferred not to say). **Panel B:** Primary includes general practitioners/family physicians, nurse practitioners/registered nurses, physician assistants and a manager of a community health center; Secondary (clinical) includes colposcopists and colposcopy registered nurses/registered practical nurses; Secondary (cytological) includes cytopathologists/technologists and pathologists; Tertiary includes gynecologists/obstetrician-gynecologists, gynecology oncologists, and gynecology nurses. **Panels B and C:** Frequency count exceeded total number of respondents as some reported multiple places of practice. DK: Don’t know; NA: Not applicable to my practice

### Answers to open-ended questions

**Table 6** presents a categorization of the open-ended feedback provided by respondents. Several topics were discerned among the diverse raw responses for each open question. Around 40% of respondents mentioned that the pandemic would facilitate HPV self-sampling and is a favorable approach to implement in cervical cancer screening. Several challenges were described including operational, implementation, and evaluation considerations as well as healthcare system considerations. Of the 206 responses to Q22, 30 survey respondents stated that they were not familiar with HPV self-sampling, whether it be with the mechanism or validity of the test. The vast majority of those who were not familiar with HPV self-sampling were primary care providers (90.0%), and the largest proportion were in Ontario (43.3%) and worked in private clinics (56.7%) **(Supplementary Figure 10)**. Similarly, 60 of the 197 respondents to Q23 explained that they were not familiar enough with HPV self-sampling to express a favorable or unfavorable opinion about its implementation as an alternative screening method. Of those, most were in Ontario (56.7%), were primary care providers (75.0%), and worked in private clinics (40.0%) **(Supplementary Figure 11)**. Respondents identified additional interactions deemed appropriate to convert to telemedicine, such as counselling services, follow-up with the patient, discussion of treatment options, and research-related activities. A substantial portion (36.2%) stated that no measures were implemented by their practice/institution to catch up with cancellations, postponements, and ongoing delays. Almost one quarter of respondents to Q52 (24.7%) were forced to interrupt the services at their practice or institution for between 2 and 4 months due to the pandemic, and 13.1% reported interruptions of over 6 months. Many of those who did not experience interruptions to their practice (11.2%) described severely reduced services, deferral of patients with lower risk or lower grade disease, and use of telemedicine. Respondents had different interpretations of Q60; whereas a few reported the continuation of regular practice (10.3%) and use of personal protective equipment (4.4%) to screen COVID-19 positive patients, most (54.4%) reported that appointments were deferred until after the patient’s isolation period. When asked about which cervical cancer screening guidelines the respondent’s practice/institution has been following, 63.5% answered governmental and 17.3% answered professional association/society.

**Table 6.**
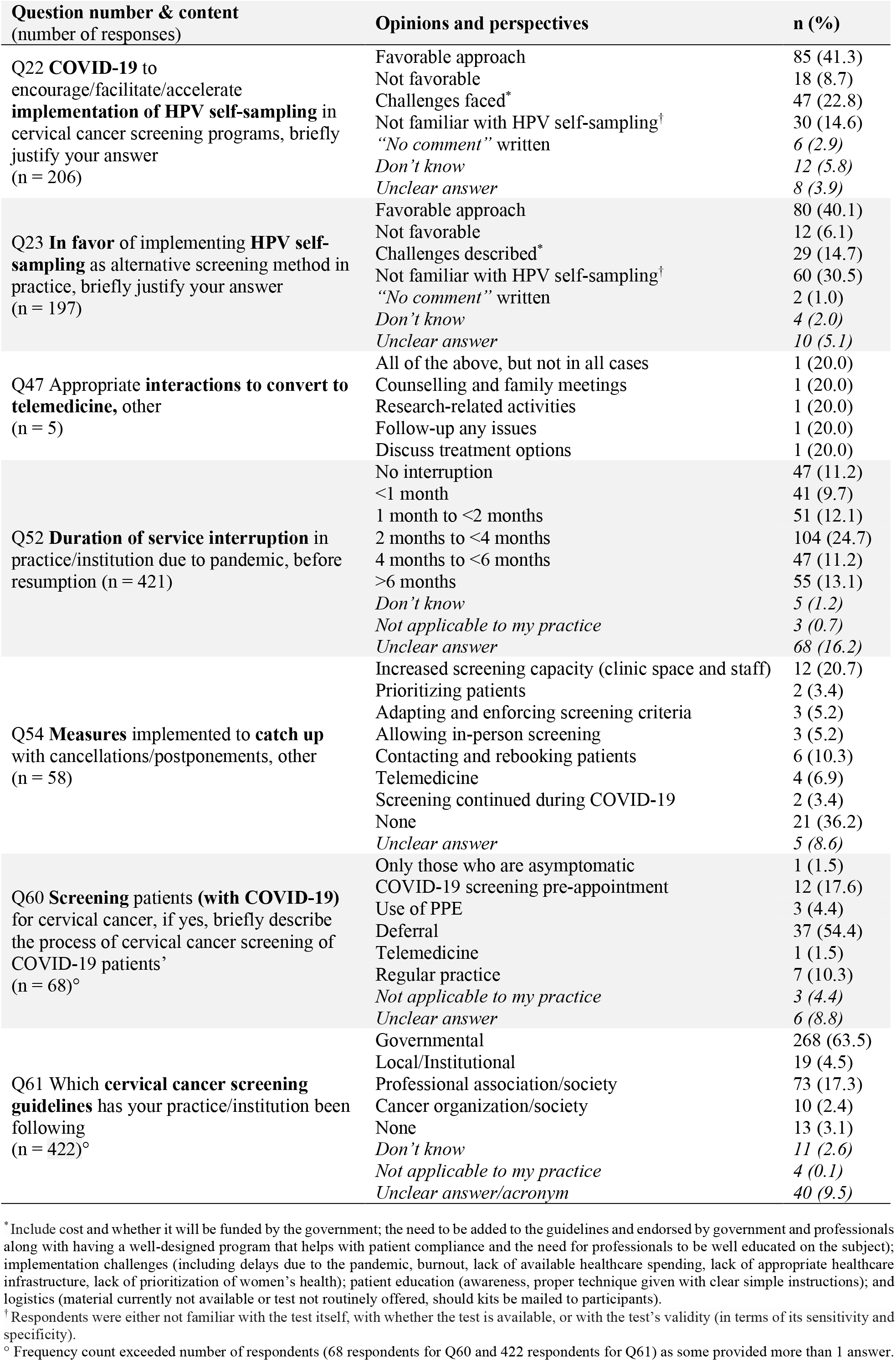
Content analysis of open-ended questions

## Discussion

We report in the current paper findings from an online Canada-wide survey of healthcare professionals, capturing their opinions, perceptions, and work experience in relation to the impact of the disruptions in routine cervical cancer screening and resulting restrictions on colposcopy services during the early period of the COVID-19 pandemic. Overall responses were reflective of the decline in cervical cancer screening and the challenges healthcare professionals faced when the pandemic was declared.

The pandemic’s negative consequences and collateral damage have been consistently observed on an international level in relation to the pauses/postponed cancer screening programs. Screening rates in the US had dropped 35% below averages of previous years (2017-2019) in the period of January-June of 2020, with an estimated 40,000 missing screening tests through March-June.^18^ A population-based study in the US reported a 46.4% decrease in the weekly number of newly identified patients with breast, colorectal, lung, pancreatic, gastric, and esophageal cancers during March-April 2020.^19^ A modelling study in the United Kingdom found that a 2-month delay in the 2-week-wait investigatory referrals for suspected cancer can lead to an estimated loss of 0-0.7 life-years per patient.^20^ In the Netherlands, a notable decrease in cancer diagnoses between February 24, 2020 and April 12, 2020 was also reported compared with the period before the COVID-19 outbreak.^6^ Decreases were observed in another study conducted in Hong Kong, where weekly colorectal cancer diagnoses had fallen by 54% during the pandemic.^21^

Particularly for cervical cancer, a two-month screening lock-down between 12 March and 8 May 2020 in Slovenia resulted in a rapid decline in screening (-92%), follow-up (-70%), and HPV triage tests (-68%), in addition to invasive diagnostic (-47%) and treatment (-15%) of cervical lesions, compared to a three-year average of years 2017-2019.^22^ An 83% decrease in the number of Pap tests was seen in Manitoba, Canada in April 2020, most likely related to limited accessibility to primary healthcare providers.^23^ During the first six months of the pandemic in Ontario, Canada, there was a decrease in the monthly average number of Pap tests (-63.8%), colposcopies (-39.7%), and treatments (-31.1%), compared with the corresponding months in 2019.^16^

Similarly, modelling data have consistently predicted an excess of cervical cancer cases and deaths caused by the scaling down of cervical screening and treatment services due to COVID-19 disruptions and resource constraints. Under a cytology-based screening model, suspensions of 6- and 24-months in the screening continuum in the US were estimated to yield an additional 5-7 and an additional 25-27 cases of cervical cancer, respectively, by 2027. The numbers of increased cases were greater for women previously screened with cytology compared with cotesting (cytology plus HPV testing).^24^ A 25.7% decrease in diagnosis of low stage cervical cancer was observed in the North of England during the pandemic compared to 2019. ^25^ The authors estimated a total of 919 cases of cervical cancer will by 2023, compared to 233 cases pre-COVID (May-October 2019), caused by a lack of diagnosis of established cases and an excess of cases caused by lack of screening. In India, delays in diagnoses and treatment were estimated to result in a 2.5% (n = 795) to 3.8% (n = 2160) lifetime increase in the deaths caused by cervical cancer, compared to no delays.^26^

Our survey results pointed to a potential for the use of self-collected samples for HPV-based screening and the need for adaptability. The World Health Organization’s call to eliminate cervical cancer^27^ has motivated efforts across the globe to scale up screening services and introduce a paradigm shift in cervical screening by implementing HPV-based programs. HPV self-sampling addresses the challenges of COVID-19 (need for social distance and a possibility of at-home sample collection) and women’s empowerment (samples collected by women themselves), thus offering a socially distanced approach that will substantially reduce the need for clinic appointments. Along the same lines, our results highlight the important role that telemedicine has played in mitigating the effects of delays in cervical cancer screening and follow-up and reducing the backlog faced upon the resumption of in-person practice.

There are some limitations to the study that need to be acknowledged. First, we were unable to fully reach out to general practitioners and family physicians who are mostly involved at the forefront of cervical cancer screening processes. The College of Family Physicians of Canada did not approve circulating the survey to their members to avoid inconveniencing them with external activities. However, we commissioned MDBriefCase to publicize the survey to community primary care providers. We also reached out to family physicians in academic medicine within university networks. Second, the non-response rate cannot be calculated due to the recruitment methods used. Finally, 34% of surveys were excluded. Most often, this was because we did not include a screening question to ensure respondents were eligible to participate. Less often, there were multiple entries by the same respondent which could have resulted from the use of a snowball method (particularly via social media) and the lack of unique IP addresses. Our survey collection strategy did not enable validation of the respondent’s eligibility to participate in the survey. However, we used the answers to the demographic and open-ended questions to determine eligibility and legitimacy of the responses and verified that there were no duplicate surveys submitted. In all, most respondents carefully answered and provided candid views.

The strengths of this survey study are its Pan-Canadian scope and design querying five themes of the cervical cancer screening and treatment continuum, the widely publicized approach and endorsements by professional societies and organizations, and the participation of multiple health professional disciplines. Its findings identified several key lessons for future response efforts and highlight the need for 1) properly formulated recommendations and strategies that would help mitigate the negative outcomes of the pandemic, 2) development of potential recovery strategies (i.e., risk-based triage systems as well as awareness campaigns on the importance and value of cervical cancer screening) for resuming routine cervical cancer screening, and 3) help building resilience in screening processes. Our survey clearly supports the implementation of HPV primary screening and the use of telemedicine to continue cervical cancer screening, treatment and follow-ups and reduce backlogs while mitigating inconveniences to both patients and healthcare professionals. In addition, insights from the survey could inform epidemiological modelling studies of the long-term effects of the interruptions and delays in screening activities on cervical cancer morbidity and mortality.

## Data Availability

Code files and datasets corresponding to analyses and descriptive figures included in the manuscript and supplement are available online at Borealis, the Canadian Dataverse Repository.

https://doi.org/10.5683/SP3/8MVU6L

## Acknowledgments

The authors are grateful to all the societies that assisted with survey distribution: Society of Canadian Colposcopists (SCC), Society of Gynecologic Oncology of Canada (GOC), Canadian Association of Pathologists (CAP), and Society of Obstetricians and Gynecologists of Canada (SOGC). We are also grateful to Dr. Wilson H Miller, Director of the COVID-19 and Cancer Program (McGill University) for the insights and support. We thank Dr. Helene Trottier (University of Montreal) for verification of the French translation of the survey questions. Finally, we express our deepest gratitude to the respondents for taking precious time out of their work schedule to share their valuable perspectives.

## Author Contributions

ELF conceptualised the survey-based study. ELF and MZ acquired funding. RA and EF formulated the survey questions under the supervision of MZ and ELF. RA and EF created the online survey. RA, MZ and ELF coordinated survey pretesting and implementation. RA, EF, and SBP verified and analyzed the data. SBP produced all tables and figures with input from MZ. MZ translated the survey to French and led the manuscript writing and tables/figures formatting. All authors were involved in data interpretation and manuscript revision.

## Data sharing

The survey instrument appears in supplementary file 1.

Upon publication, the survey dataset will be deposited in the McGill Dataverse repository (https://dataverse.scholarsportal.info/dataverse/mcgill).

## Notes

**Funding:** The present work was supported by the Canadian Institutes of Health Research (operating grant COVID-19 May 2020 Rapid Research Funding Opportunity VR5-172666 Rapid Research competition and foundation grant 143347 to Eduardo L. Franco). Eliya Farah and Rami Ali each received a MSc. stipend from the Department of Oncology, McGill University.

**Competing Interests:** The authors have no conflicts of interest to disclose related to the work in this manuscript.

### Competing Interest Statement

All authors have completed the ICMJE uniform disclosure form at www.icmje.org/coi_disclosure.pdf and declare: MZ and SBP have no conflicts of interest to disclose. RA and EF received MSc. Stipends from the Gerald Bronfman Department of Oncology, McGill University. ELF reports support for the present manuscript in the form of a grant to his institution is his name from the Canadian Institutes of Health Research and the Cancer Research Society; consultancy for Merck; a patent related to the discovery "DNA methylation markers for early detection of cervical cancer", registered at the Office of Innovation and Partnerships, McGill University, Montreal, Quebec, Canada; and financial interests with Elsevier and Elifesciences Ltd in the form of support fees to maintain the editorial office and work as Senior Editor, respectively. No other relationships or activities could appear to have influenced the submitted work.

### Funding Statement

The present work was supported by the Canadian Institutes of Health Research (CIHR-COVID-19 Rapid Research Funding opportunity, VR5-172666 grant to Eduardo L. Franco). Eliya Farah and Rami Ali each received MSc. stipends from the Gerald Bronfman Department of Oncology, McGill University.

### Author Declarations

The Faculty of Medicine Institutional Review Board (IRB) of McGill University granted ethical approval for this work.

## References

1. Ghebreyesus TA. WHO Director-General’s opening remarks at the media briefing on COVID-19 - 11 March 2020. WHO Director General’s speeches. https://www.who.int/director-general/speeches/detail/who-director-general-s-opening-remarks-at-the-media-briefing-on-covid-19 11-march-2020. Published 2020. Accessed February 25, 2022.

2. Blumenthal D, Fowler EJ, Abrams M, Collins SR. Sounding Board: Covid-19 - Implications for the Health Care System. N Engl J Med. 2020;383(15):1483–1488. doi:10.1056/NEJMsb2021088

3. McMahon M, Thompson E, Nadigel J, Glazier RH. Informing Canada’s health system response to COVID-19: Priorities for health services and policy research. Healthc Policy. 2020;16(1):112–124. doi:10.12927/hcpol.2020.26249

4. Farah E, Ali R, Tope P, et al. A review of Canadian cancer-related clinical practice guidelines and resources during the covid-19 pandemic. Curr Oncol. 2021;28(2):1020–1033. doi:10.3390/curroncol28020100

5. Price ST, Mainous AG, Rooks BJ. Survey of cancer screening practices and telehealth services among primary care physicians during the COVID-19 pandemic. Prev Med Reports. 2022;27:101769. doi:10.1016/j.pmedr.2022.101769

6. Dinmohamed AG, Visser O, Verhoeven RHA, et al. Fewer cancer diagnoses during the COVID-19 epidemic in the Netherlands. Lancet Oncol. 2020;21(6):750–751. doi:10.1016/S1470-2045(20)30265-5

7. Cancer Australia. Review of the Impact of COVID-19 on Medical Services and Procedures in Australia Utilising MBS Data: Lung and Prostate Cancers. Surry Hills, NSW: Cancer Australia; 2020. https://www.canceraustralia.gov.au/publications-and-resources/cancer-australia-publications/review-impact-covid-19-medical-services-and-procedures-australia-utilising-mbs-data.

8. Gourd E. COVID-19 pandemic causes cervical cancer screening crisis. Lancet Oncol. 2021;22(8):1060. doi:10.1016/S1470-2045(21)00382-X

9. Wilkinson E. How cancer services are fighting to counter covid-19’s impact. BMJ. 2020;370:10–12. doi:10.1136/bmj.m2747

10. Maringe C, Spicer J, Morris M, et al. The impact of the COVID-19 pandemic on cancer deaths due to delays in diagnosis in England, UK: a national, population-based, modelling study. Lancet Oncol. 2020. doi:10.1016/s1470-2045(20)30388-0

11. Chen-See S. Disruption of cancer care in Canada during COVID-19. Lancet Oncol. 2020;21(8):e374. doi:10.1016/S1470-2045(20)30397-1

12. Canadian Cancer Survivor Network. New survey reveals cancer care disruption in Canada has triggered another public health crisis. Canadian Cancer Survivor Network. https://survivornet.ca/news/cancer-patients-face-double-jeopardy-with-covid-19/. Published 2020. Accessed March 2, 2022.

13. Malagón T, Yong JHE, Tope P, et al. Predicted long-term impact of COVID-19 pandemic-related care delays on cancer mortality in Canada. Int J Cancer. 2021;150:1244–1254. doi:10.1002/ijc.33884

14. DeGroff A, Miller J, Sharma K, et al. COVID-19 impact on screening test volume through the National Breast and Cervical Cancer early detection program, January-June 2020, in the United States. Prev Med (Baltim). 2021;151:106559. doi:10.1016/j.ypmed.2021.106559

15. Walker MJ, Meggetto O, Gao J, et al. Measuring the impact of the COVID-19 pandemic on organized cancer screening and diagnostic follow-up care in Ontario, Canada: A provincial, population-based study. Prev Med (Baltim). 2021;151:106586. doi:10.1016/j.ypmed.2021.106586

16. Meggetto O, Jembere N, Gao J, et al. The impact of the COVID-19 pandemic on the Ontario Cervical Screening Program, colposcopy and treatment services in Ontario, Canada: a population-based study. BJOG An Int J Obstet Gynaecol. 2021;128(9):1503–1510. doi:10.1111/1471-0528.16741

17. Eysenbach G. Improving the quality of web surveys: The Checklist for Reporting Results of Internet E-Surveys (CHERRIES). J Med Internet Res. 2004;6(3):e34. doi:10.2196/jmir.6.3.e34

18. Cox C, Amin K, Rae M, Gallagher K. Early 2021 Data Show No Rebound in Health Care Utilization.; 2021.

19. Kaufman HW, Chen Z, Niles JK, Fesko YA. Changes in Newly Identified Cancer among US Patients from before COVID-19 through the First Full Year of the Pandemic. JAMA Netw Open. 2021. doi:10.1001/jamanetworkopen.2021.25681

20. Sud A, Torr B, Jones ME, et al. Effect of delays in the 2-week-wait cancer referral pathway during the COVID-19 pandemic on cancer survival in the UK: a modelling study. Lancet Oncol. 2020;21(8):1035–1044. doi:10.1016/S1470-2045(20)30392-2

21. Lui TKL, Leung K, Guo CG, Tsui VWM, Wu JT, Leung WK. Impacts of the Coronavirus 2019 Pandemic on Gastrointestinal Endoscopy Volume and Diagnosis of Gastric and Colorectal Cancers: A Population-Based Study. Gastroenterology. 2020;159(3):1164-1166.e3. doi:10.1053/j.gastro.2020.05.037

22. Ivanuš U, Jerman T, Gašper Oblak U, et al. The impact of the COVID-19 pandemic on organised cervical cancer screening: The first results of the Slovenian cervical screening programme and registry. Lancet Reg Heal - Eur. 2021;5:100101. doi:10.1016/j.lanepe.2021.100101

23. Decker KM, Feely A, Bucher O, Singh H, Turner D, Lambert P. Evaluating the impact of the COVID-19 pandemic on cancer screening in a central Canadian province. Prev Med (Baltim). 2022;155:106961. doi:10.1016/j.ypmed.2022.106961

24. Burger EA, Jansen EEL, Killen J, et al. Impact of COVID-19-related care disruptions on cervical cancer screening in the United States. J Med Screen. 2021. doi:10.1177/09691413211001097

25. Davies JM, Spencer A, Macdonald S, et al. Cervical cancer and COVID—an assessment of the initial effect of the pandemic and subsequent projection of impact for women in England: A cohort study. BJOG An Int J Obstet Gynaecol. June 2022. doi:10.1111/1471-0528.17098

26. Gupta N, Akashdeep ;, Chauhan S, Shankar Prinja ;, Pandey AK. Impact of COVID-19 on Outcomes for Patients With Cervical Cancer in India. JCO Glob Oncol. 2021;7:716–725. https://ascopubs.org/go/authors/open-access.

