## Supplemental Tables 1-9, Supplemental Figures 1-11 for "Pan-Canadian survey on the impact of the COVID-19 pandemic on cervical cancer screening and management"

### Supplementary Tables

#### Theme 1: Screening practice

**STable 1.** Pandemic-related updates to practice/institutional/jurisdictional procedures (n = 510)

| Question number & content | Categories | n (%) |
|---|---|---|
| Q11 Allowance of **in-person consultations** during peak pandemic | Yes | 378 (74.1) |
| | No | 99 (19.4) |
| | *Don't know* | *13 (2.6)* |
| | *Not applicable to my practice* | *20 (3.9)* |
| Q12 Informed about **latest recommendations/guidelines** for cervical cancer screening during peak pandemic | Yes | 274 (53.7) |
| | No | 164 (32.2) |
| | *Don't know* | *62 (12.2)* |
| | *Not applicable to my practice* | *10 (2.0)* |
| Q13 **Frequency of updates** to recommendations/guidelines that modify scheduling/performing cervical cancer screening | Every week | 18 (3.5) |
| | Every 2 weeks | 48 (9.4) |
| | Every month | 101 (19.8) |
| | Every 2 months | 58 (11.4) |
| | >2 months | 118 (23.1) |
| | Never | 92 (18.0) |
| | *Don't know* | *65 (12.8)* |
| | *Not applicable to my practice* | *10 (2.0)* |

**STable 2.** Screening test practices (n = 469)

| Question number & content | | Categories | n (%) |
|---|---|---|---|
| Q14 **Type of test** used for primary cervical cancer screening | **Pap** test | Yes | 357 (76.1) |
| | | No | 61 (13.0) |
| | | *Don't know* | *37 (7.9)* |
| | | *Not applicable to my practice* | *14 (3.0)* |
| | **HPV** test | Yes | 154 (32.8) |
| | | No | 230 (49.0) |
| | | *Don't know* | *36 (7.7)* |
| | | *Not applicable to my practice* | *49 (10.5)* |
| | **HPV/Pap** co-test | Yes | 119 (25.4) |
| | | No | 262 (55.9) |
| | | *Don't know* | 43 (9.2) |
| | | *Not applicable to my practice* | 45 (9.6) |
| **Q15 Change in number of screening tests** compared to pre-COVID-19 | **Pap** test | Decrease ≥75% | 74 (15.8) |
| | | Decrease 26-74% | 117 (25.0) |
| | | Decrease ≤25% | 95 (20.3) |
| | | Unaffected | 82 (17.5) |
| | | Increase ≤25% | 24 (5.1) |
| | | Increase 26-74% | 18 (3.8) |
| | | Increase ≥75% | 9 (1.9) |
| | | *Don't know* | *37 (7.9)* |
| | | *Not applicable to my practice* | *13 (2.8)* |
| | **HPV** test | Decrease ≥75% | 23 (4.9) |
| | | Decrease 26-74% | 31 (6.6) |
| | | Decrease ≤25% | 33 (7.0) |
| | | Unaffected | 91 (19.4) |
| | | Increase ≤25% | 34 (7.3) |
| | | Increase 26-74% | 30 (6.4) |
| | | Increase ≥75% | 11 (2.4) |
| | | *Don't know* | *43 (9.2)* |
| | | *Not applicable to my practice* | *173 (37.0)* |
| | **HPV/Pap** co-test | Decrease ≥75% | 18 (3.8) |
| | | Decrease 26-74% | 36 (7.7) |
| | | Decrease ≤25% | 35 (7.5) |
| | | Unaffected | 70 (14.9) |
| | | Increase ≤25% | 36 (7.7) |
| | | Increase 26-74% | 22 (4.7) |
| | | Increase ≥75% | 12 (2.6) |
| | | *Don't know* | *39 (8.3)* |
| | | *Not applicable to my practice* | *201 (42.9)* |
| **Q16 Delays** in scheduling of **tests** | **Pap** test | Yes | 267 (56.9) |
| | | No | 132 (28.1) |
| | | *Don't know* | *50 (10.7)* |
| | | *Not applicable to my practice* | *20 (4.3)* |
| | **HPV** test | Yes | 106 (22.6) |
| | | No | 116 (24.7) |
| | | *Don't know* | 57 (12.2) |
| | | *Not applicable to my practice* | 190 (40.5) |
| | **HPV/Pap** co-test | Yes | 102 (21.8) |
| | | No | 93 (19.8) |
| | | *Don't know* | *58 (12.4)* |
| | | *Not applicable to my practice* | *216 (46.1)* |

HPV: human papillomavirus

**STable 3.** Cancellation and postponement of scheduled screening tests by type of test

| **Question number & content** (number of responses) | | **Categories** | **n (%)** |
|---|---|---|---|
| Q17 Percentage of **scheduled screening tests** that have been **cancelled** (n = 469) | **Pap** test | 0% | 76 (16.2) |
| | | 1-24% | 145 (31.0) |
| | | 25-49% | 80 (17.1) |
| | | 50-74% | 45 (9.6) |
| | | ≥75% | 49 (10.5) |
| | | *Don't know* | *54 (11.5)* |
| | | *Not applicable to my practice* | *20 (4.3)* |
| | **HPV** test | 0% | 65 (13.9) |
| | | 1-24% | 41 (8.7) |
| | | 25-49% | 52 (11.1) |
| | | 50-74% | 33 (7.0) |
| | | ≥75% | 37 (7.9) |
| | | *Don't know* | *56 (11.9)* |
| | | *Not applicable to my practice* | *185 (39.5)* |
| | **HPV/Pap** co-test | 0% | 49 (10.5) |
| | | 1-24% | 43 (9.2) |
| | | 25-49% | 38 (8.1) |
| | | 50-74% | 39 (8.3) |
| | | ≥75% | 42 (9.0) |
| | | *Don't know* | *45 (9.6)* |
| | | *Not applicable to my practice* | *213 (45.4)* |
| Q18 Percentage of **scheduled screening tests** that have been **postponed** (n = 468) | **Pap** test | 0% | 29 (6.2) |
| | | 1-24% | 119 (25.4) |
| | | 25-49% | 100 (21.4) |
| | | 50-74% | 75 (16.0) |
| | | ≥75% | 78 (16.7) |
| | | *Don't know* | *48 (10.3)* |
| | | *Not applicable to my practice* | *19 (4.1)* |
| | **HPV** test | 0% | 49 (10.5) |
| | | 1-24% | 49 (10.5) |
| | | 25-49% | 55 (11.8) |
| | | 50-74% | 36 (7.7) |
| | | ≥75% | 45 (9.6) |
| | | *Don't know* | *47 (10.0)* |
| | | *Not applicable to my practice* | *187 (40.0)* |
| | **HPV/Pap** co-test | 0% | 44 (9.4) |
| | | 1-24% | 45 (9.6) |
| | | 25-49% | 49 (10.5) |
| | | 50-74% | 43 (9.2) |
| | | ≥75% | 32 (6.8) |
| | | *Don't know* | *45 (9.6)* |
| | | *Not applicable to my practice* | *210 (44.9)* |
| | **Pap** test | 1 week to <2 weeks | 15 (3.2) |
| | | 2 weeks to <4 weeks | 43 (9.2) |
| | | 1 month to <2 months | 78 (16.7) |
| | | 2 months to <4 months | 128 (27.4) |
| | | 4 months to <6 months | 87 (18.7) |
| | | >6 months | 47 (10.1) |
| | | *Don't know* | 36 (7.7) |
| | | *Not applicable to my practice* | 33 (7.1) |
| | | 1 week to <2 weeks | 22 (4.7) |
| | | 2 weeks to <4 weeks | 31 (6.6) |
| | | 1 month to <2 months | 46 (9.9) |

| **Question number & content** (number of responses) | | Categories | n (%) |
|---|---|---|---|
| **Q19 Deferral** period of postponed **testing appointments** (n = 467) | **HPV** test | 2 months to <4 months | 48 (10.3) |
| | | 4 months to <6 months | 35 (7.5) |
| | | >6 months | 31 (6.6) |
| | | *Don't know* | 42 (9.0) |
| | | *Not applicable to my practice* | 212 (45.4) |
| | **HPV/Pap** co-test | 1 week to <2 weeks | 16 (3.4) |
| | | 2 weeks to <4 weeks | 27 (5.8) |
| | | 1 month to <2 months | 30 (6.4) |
| | | 2 months to <4 months | 46 (9.9) |
| | | 4 months to <6 months | 47 (10.1) |
| | | >6 months | 31 (6.6) |
| | | *Don't know* | 36 (7.7) |
| | | *Not applicable to my practice* | 234 (50.1) |

HPV: human papillomavirus

**STable 4.** Pandemic-related delays in forwarding screening test samples to the lab (n = 467)

| Question number & content | | Categories | n (%) |
|---|---|---|---|
| **Q20 Delay** in **forwarding tests to labs** for processing | **Pap** test | Yes | 70 (15.0) |
| | | No | 323 (69.2) |
| | | *Don't know* | *57 (12.2)* |
| | | *Not applicable to my practice* | *17 (3.6)* |
| | **HPV** test | Yes | 73 (15.6) |
| | | No | 164 (35.1) |
| | | *Don't know* | *42 (9.0)* |
| | | *Not applicable to my practice* | *188 (40.3)* |
| | **HPV/Pap** co-test | Yes | 65 (13.9) |
| | | No | 126 (27.0) |
| | | *Don't know* | *58 (12.4)* |
| | | *Not applicable to my practice* | *218 (46.7)* |
| **Q21 Delay** period in **forwarding test** samples to lab | **Pap** test | 1 week to <2 weeks | 73 (15.6) |
| | | 2 weeks to <4 weeks | 32 (6.9) |
| | | 1 month to <2 months | 45 (9.6) |
| | | 2 months to <4 months | 39 (8.4) |
| | | 4 months to <6 months | 24 (5.1) |
| | | >6 months | 6 (1.3) |
| | | *Don't know* | *63 (13.5)* |
| | | *Not applicable to my practice* | *185 (39.6)* |
| | **HPV** test | 1 week to <2 weeks | 35 (7.5) |
| | | 2 weeks to <4 weeks | 29 (6.2) |
| | | 1 month to <2 months | 32 (6.9) |
| | | 2 months to <4 months | 28 (6.0) |
| | | 4 months to <6 months | 34 (7.3) |
| | | >6 months | 22 (4.7) |
| | | *Don't know* | *40 (8.6)* |
| | | *Not applicable to my practice* | *247 (52.9)* |
| | **HPV/Pap** co-test | 1 week to <2 weeks | 32 (6.9) |
| | | 2 weeks to <4 weeks | 25 (5.4) |
| | | 1 month to <2 months | 24 (5.1) |
| | | 2 months to <4 months | 37 (7.9) |
| | | 4 months to <6 months | 27 (5.8) |
| | | >6 months | 18 (3.9) |
| | | *Don't know* | *43 (9.2)* |
| | | *Not applicable to my practice* | *261 (55.9)* |

HPV: human papillomavirus

**STable 5.** Professional opinions on HPV self-sampling (n = 455)

| **Question number & content** | **Categories** | **n (%)** |
|---|---|---|
| Q22 **COVID-19** to encourage/facilitate/accelerate **implementation of HPV self-sampling** in cervical cancer screening programs | Yes | 150 (33.0) |
| | No | 143 (31.4) |
| | Maybe | 162 (35.6) |
| Q23 **In favor** of implementing **HPV self-sampling** as alternative screening method in practice | Yes | 228 (50.1) |
| | No | 123 (27.0) |
| | Maybe | 104 (22.9) |

HPV: human papillomavirus

**STable 6.** Changes in colposcopy appointments

| **Question number & content** (number of responses) | | **Categories** | **n (%)** |
|---|---|---|---|
| Q24 **Changes in number of colposcopy-biopsy** procedures compared to pre-COVID-19 (n = 452) | | Decrease ≥75% | 21 (4.7) |
| | | Decrease 26-74% | 61 (13.5) |
| | | Decrease ≤25% | 87 (19.3) |
| | | Unaffected | 40 (8.9) |
| | | Increase ≤25% | 24 (5.3) |
| | | Increase 26-74% | 10 (2.2) |
| | | Increase ≥75% | 6 (1.3) |
| | | *Don't know* | *34 (7.5)* |
| | | *Not applicable to my practice* | *169 (37.4)* |
| Q25 **Cancellations** of **colposcopy** appointments (n = 452) | | Yes | 114 (25.2) |
| | | No | 95 (21.0) |
| | | *Don't know* | *72 (15.9)* |
| | | *Not applicable to my practice* | *171 (37.8)* |
| Q26 Percentage of **cancelled colposcopy** appointments (n = 114) | Cancelled by **physician or provider's institution** | 0% | 5 (4.4) |
| | | 1-24% | 45 (39.5) |
| | | 25-49% | 30 (26.3) |
| | | 50-74% | 16 (14.0) |
| | | ≥75% | 11 (9.7) |
| | | *Don't know* | *7 (6.1)* |
| | Cancelled by **patient** | 0% | 6 (5.3) |
| | | 1-24% | 41 (36.0) |
| | | 25-49% | 23 (20.2) |
| | | 50-74% | 21 (18.4) |
| | | ≥75% | 14 (12.3) |
| | | *Don't know* | *9 (7.9)* |
| Q27 **Postponements** in scheduling of **colposcopy appointments** (n = 452) | | Yes | 167 (37.0) |
| | | No | 68 (15.0) |
| | | *Don't know* | *44 (9.7)* |
| | | *Not applicable to my practice* | *173 (38.3)* |
| Q28 Percentage of **postponed colposcopy** appointments (n = 167) | Postponed by **physician or provider's institution** | 0% | 2 (1.2) |
| | | 1-24% | 47 (28.1) |
| | | 25-49% | 40 (24.0) |
| | | 50-74% | 43 (25.8) |
| | | ≥75% | 25 (15.0) |
| | | *Don't know* | *10 (6.0)* |
| | Postponed by **patient** | 0% | 7 (4.2) |
| | | 1-24% | 62 (37.1) |
| | | 25-49% | 44 (26.4) |
| | | 50-74% | 23 (13.8) |
| | | ≥75% | 14 (8.4) |
| | | *Don't know* | *17 (10.2)* |
| **Q29 Length of deferral** period of postponed **colposcopy appointments** (n = 167) | | 1 week to <2 weeks | 6 (3.6) |
| | | 2 weeks to <4 weeks | 18 (10.8) |
| | | 1 month to <2 months | 44 (26.4) |
| | | 2 months to <4 months | 59 (35.3) |
| | | 4 months to <6 months | 25 (15.0) |
| | | >6 months | 9 (5.4) |
| | | *Don't know* | *6 (3.6)* |

**STable 7.** Changes in follow-up appointments

| **Question number & content** (number of responses) | | **Categories** | **n (%)** |
|---|---|---|---|
| Q30 **Delay in follow-up** of patients following test results (n = 446) | Abnormal cytology | Yes | 121 (27.1) |
| | | No | 251 (56.3) |
| | | *Don't know* | 52 (11.7) |
| | | *Not applicable to my practice* | 22 (4.9) |
| | High-grade lesions | Yes | 97 (21.8) |
| | | No | 257 (57.6) |
| | | *Don't know* | 57 (12.8) |
| | | *Not applicable to my practice* | 35 (7.9) |
| | Positive HPV test | Yes | 75 (16.8) |
| | | No | 191 (42.8) |
| | | *Don't know* | 58 (13.0) |
| | | *Not applicable to my practice* | 122 (27.4) |
| | Positive HPV self-sampling | Yes | 55 (12.3) |
| | | No | 96 (21.5) |
| | | *Don't know* | 52 (11.7) |
| | | *Not applicable to my practice* | 243 (54.5) |
| Q31 **Cancellations** of **follow-up** appointments (n = 445) | | Yes | 148 (33.3) |
| | | No | 170 (38.2) |
| | | *Don't know* | 97 (21.8) |
| | | *Not applicable to my practice* | 30 (6.7) |
| Q32 Percentage of **cancelled follow-up** appointments (n = 148) | Cancelled by **physician or provider's institution** | 0% | 25 (16.9) |
| | | 1-24% | 58 (39.2) |
| | | 25-49% | 29 (19.6) |
| | | 50-74% | 19 (12.8) |
| | | ≥75% | 9 (6.1) |
| | | *Don't know* | 8 (5.4) |
| | Cancelled by **patient** | 0% | 3 (2.0) |
| | | 1-24% | 61 (41.2) |
| | | 25-49% | 35 (23.7) |
| | | 50-74% | 28 (18.9) |
| | | ≥75% | 15 (10.1) |
| | | *Don't know* | 6 (4.1) |
| | **Converted to telemedicine** | 0% | 24 (16.2) |
| | | 1-24% | 44 (29.7) |
| | | 25-49% | 29 (19.6) |
| | | 50-74% | 26 (17.6) |
| | | ≥75% | 14 (9.5) |
| | | *Don't know* | 11 (7.4) |
| **Q33 Postponements** in scheduling of **follow-up appointments** (n = 445) | | Yes | 238 (53.5) |
| | | No | 123 (27.6) |
| | | *Don't know* | 55 (12.4) |
| | | *Not applicable to my practice* | 29 (6.5) |
| Q34 Percentage of **postponed follow-up** appointments (n=238) | Postponed by **physician or provider's institution** | 0% | 21 (8.8) |
| | | 1-24% | 84 (35.3) |
| | | 25-49% | 49 (20.6) |
| | | 50-74% | 40 (16.8) |
| | | ≥75% | 28 (11.8) |
| | | *Don't know* | *16 (6.7)* |
| | Postponed by **patient** | 0% | 6 (2.5) |
| | | 1-24% | 98 (41.2) |
| | | 25-49% | 59 (24.8) |
| | | 50-74% | 36 (15.1) |

| | | | |
|---|---|---|---|
| | | ≥75% | 19 (8.0) |
| | | *Don't know* | *20 (8.4)* |
| | **Converted to telemedicine** | 0% | 48 (20.2) |
| | | 1-24% | 51 (21.4) |
| | | 25-49% | 41 (17.2) |
| | | 50-74% | 42 (17.6) |
| | | ≥75% | 29 (12.2) |
| | | *Don't know* | *27 (11.3)* |
| **Q35 Length of deferral** period of postponed **follow-up appointments** (n=238) | | 1 week to <2 weeks | 11 (4.6) |
| | | 2 weeks to <4 weeks | 39 (16.4) |
| | | 1 month to <2 months | 67 (28.2) |
| | | 2 months to <4 months | 69 (29.0) |
| | | 4 months to <6 months | 35 (14.7) |
| | | >6 months | 9 (3.8) |
| | | *Don't know* | *8 (3.4)* |

HPV: human papillomavirus

**STable 8.** Pandemic-related delays in receiving test results from lab (n = 445)

| **Question number & content** | | **Categories** | **n (%)** |
|---|---|---|---|
| Q36 **Delay in receiving test results** from lab | **Pap** test | Yes | 94 (21.1) |
| | | No | 273 (61.4) |
| | | *Don't know* | 65 (14.6) |
| | | *Not applicable to my practice* | 13 (2.9) |
| | **HPV** test | Yes | 86 (19.3) |
| | | No | 130 (29.2) |
| | | *Don't know* | 63 (14.2) |
| | | *Not applicable to my practice* | 166 (37.3) |
| | **HPV/Pap** co-test | Yes | 54 (12.1) |
| | | No | 116 (26.1) |
| | | *Don't know* | 76 (17.1) |
| | | *Not applicable to my practice* | 199 (44.7) |
| **Q37 Delay** period in **receiving test** samples from lab | **Pap** test | 1 week to <2 weeks | 62 (13.9) |
| | | 2 weeks to <4 weeks | 49 (11.0) |
| | | 1 month to <2 months | 58 (13.0) |
| | | 2 months to <4 months | 45 (10.1) |
| | | 4 months to <6 months | 28 (6.3) |
| | | >6 months | 15 (3.4) |
| | | *Don't know* | 56 (12.6) |
| | | *Not applicable to my practice* | 132 (29.7) |
| | **HPV** test | 1 week to <2 weeks | 30 (6.8) |
| | | 2 weeks to <4 weeks | 31 (7.0) |
| | | 1 month to <2 months | 37 (8.3) |
| | | 2 months to <4 months | 35 (7.9) |
| | | 4 months to <6 months | 35 (7.9) |
| | | >6 months | 17 (3.8) |
| | | *Don't know* | 42 (9.4) |
| | | *Not applicable to my practice* | 218 (49.0) |
| | **HPV/Pap** co-test | 1 week to <2 weeks | 29 (6.5) |
| | | 2 weeks to <4 weeks | 23 (5.2) |
| | | 1 month to <2 months | 28 (6.3) |
| | | 2 months to <4 months | 32 (7.2) |
| | | 4 months to <6 months | 31 (7.0) |
| | | >6 months | 19 (4.3) |
| | | *Don't know* | 51 (11.5) |
| | | *Not applicable to my practice* | 232 (52.1) |

HPV: human papillomavirus

### Theme 2: Treatment of pre-cancerous lesions and cancer

**STable 9.** Changes observed in number of treatment procedures, by treatment type (n = 431)

| Variable | | Categories | n (%) |
|---|---|---|---|
| Q38 Changes in **number of treatment procedures** | Cold knife conization | Decrease ≥75% | 14 (3.3) |
| | | Decrease 26-74% | 25 (5.8) |
| | | Decrease ≤25% | 29 (6.7) |
| | | Unaffected | 54 (12.5) |
| | | Increase ≤25% | 28 (6.5) |
| | | Increase 26-74% | 27 (6.3) |
| | | Increase ≥75% | 13 (3.0) |
| | | *Don't know* | *41 (9.5)* |
| | | *Not applicable to my practice* | *200 (46.4)* |
| | Other excisional | Decrease ≥75% | 14 (3.3) |
| | | Decrease 26-74% | 32 (7.4) |
| | | Decrease ≤25% | 40 (9.3) |
| | | Unaffected | 55 (12.8) |
| | | Increase ≤25% | 32 (7.4) |
| | | Increase 26-74% | 23 (5.3) |
| | | Increase ≥75% | 12 (2.8) |
| | | *Don't know* | *34 (7.9)* |
| | | *Not applicable to my practice* | *189 (43.9)* |
| | Ablative procedures | Decrease ≥75% | 15 (3.5) |
| | | Decrease 26-74% | 24 (5.6) |
| | | Decrease ≤25% | 29 (6.7) |
| | | Unaffected | 47 (10.9) |
| | | Increase ≤25% | 31 (7.2) |
| | | Increase 26-74% | 24 (5.6) |
| | | Increase ≥75% | 15 (3.5) |
| | | *Don't know* | *36 (8.4)* |
| | | *Not applicable to my practice* | *210 (48.7)* |
| | Hysterectomy | Decrease ≥75% | 20 (4.6) |
| | | Decrease 26-74% | 36 (8.4) |
| | | Decrease ≤25% | 47 (10.9) |
| | | Unaffected | 40 (9.3) |
| | | Increase ≤25% | 26 (6.0) |
| | | Increase 26-74% | 19 (4.4) |
| | | Increase ≥75% | 13 (3.0) |
| | | *Don't know* | *37 (8.6)* |
| | | *Not applicable to my practice* | *193 (44.8)* |
| | Chemotherapy | Decrease ≥75% | 6 (1.4) |
| | | Decrease 26-74% | 12 (2.8) |
| | | Decrease ≤25% | 29 (6.7) |
| | | Unaffected | 47 (10.9) |
| | | Increase ≤25% | 30 (7.0) |
| | | Increase 26-74% | 19 (4.4) |
| | | Increase ≥75% | 16 (3.7) |
| | | *Don't know* | *40 (9.3)* |
| | | *Not applicable to my practice* | *232 (53.8)* |
| | Radiation | Decrease ≥75% | 7 (1.6) |
| | | Decrease 26-74% | 13 (3.0) |
| | | Decrease ≤25% | 39 (9.1) |
| | | Unaffected | 34 (7.9) |
| | | Increase ≤25% | 27 (6.3) |

| | | | |
|---|---|---|---|
| | | Increase 26-74% | 21 (4.9) |
| | | Increase ≥75% | 12 (2.8) |
| | | *Don't know* | *42 (9.7)* |
| | | *Not applicable to my practice* | *236 (54.8)* |
| Q39 **Cancellations / postponements of treatment procedures** | Cold knife conization | Yes | 58 (13.5) |
| | | No | 99 (23.0) |
| | | *Don't know* | *63 (14.6)* |
| | | *Not applicable to my practice* | *211 (49.0)* |
| | Other excisional | Yes | 102 (23.7) |
| | | No | 78 (18.1) |
| | | *Don't know* | *62 (14.4)* |
| | | *Not applicable to my practice* | *188 (43.9)* |
| | Ablative procedures | Yes | 84 (19.5) |
| | | No | 81 (18.8) |
| | | *Don't know* | *54 (12.5)* |
| | | *Not applicable to my practice* | *212 (49.2)* |
| | Hysterectomy | Yes | 91 (21.1) |
| | | No | 79 (18.3) |
| | | *Don't know* | *61 (14.2)* |
| | | *Not applicable to my practice* | *200 (46.4)* |
| | Chemotherapy | Yes | 45 (10.4) |
| | | No | 82 (19.0) |
| | | *Don't know* | *69 (16.0)* |
| | | *Not applicable to my practice* | *235 (54.5)* |
| | Radiation | Yes | 50 (11.6) |
| | | No | 72 (16.7) |
| | | *Don't know* | *72 (16.7)* |
| | | *Not applicable to my practice* | *237 (55.0)* |
| Q40 Percentage of **scheduled treatment procedures** that were **cancelled** | Cold knife conization | 0% | 37 (8.6) |
| | | 1-24% | 34 (7.9) |
| | | 25-49% | 47 (10.9) |
| | | 50-74% | 29 (6.7) |
| | | ≥75% | 29 (6.7) |
| | | *Don't know* | *52 (12.1)* |
| | | *Not applicable to my practice* | *203 (47.3)* |
| | Other excisional | 0% | 44 (10.2) |
| | | 1-24% | 43 (10.0) |
| | | 25-49% | 47 (10.9) |
| | | 50-74% | 40 (9.3) |
| | | ≥75% | 22 (5.1) |
| | | *Don't know* | *50 (11.6)* |
| | | *Not applicable to my practice* | *185 (42.9)* |
| | Ablative procedures | 0% | 35 (8.1) |
| | | 1-24% | 22 (5.1) |
| | | 25-49% | 49 (11.4) |
| | | 50-74% | 31 (7.2) |
| | | ≥75% | 30 (7.0) |
| | | *Don't know* | *59 (13.7)* |
| | | *Not applicable to my practice* | *205 (47.6)* |
| | Hysterectomy | 0% | 40 (9.3) |
| | | 1-24% | 36 (8.4) |
| | | 25-49% | 40 (9.3) |
| | | 50-74% | 35 (8.1) |
| | | ≥75% | 31 (7.2) |
| | | *Don't know* | *54 (12.5)* |

| | | | |
|---|---|---|---|
| | | *Not applicable to my practice* | *195 (45.2)* |
| | Chemotherapy | 0% | 20 (4.7) |
| | | 1-24% | 36 (8.4) |
| | | 25-49% | 36 (8.4) |
| | | 50-74% | 34 (7.9) |
| | | ≥75% | 20 (4.6) |
| | | *Don't know* | *57 (13.2)* |
| | | *Not applicable to my practice* | *228 (52.9)* |
| | Radiation | 0% | 21 (4.9) |
| | | 1-24% | 33 (7.7) |
| | | 25-49% | 36 (8.4) |
| | | 50-74% | 32 (7.4) |
| | | ≥75% | 26 (6.0) |
| | | *Don't know* | *53 (12.3)* |
| | | *Not applicable to my practice* | *230 (53.4)* |
| Q41 Percentage of **scheduled treatment procedures** that were **postponed** | Cold knife conization | 0% | 26 (6.0) |
| | | 1-24% | 46 (10.7) |
| | | 25-49% | 46 (10.7) |
| | | 50-74% | 37 (8.6) |
| | | ≥75% | 20 (4.7) |
| | | *Don't know* | *52 (12.1)* |
| | | *Not applicable to my practice* | *204 (47.3)* |
| | Other excisional | 0% | 24 (5.6) |
| | | 1-24% | 57 (13.2) |
| | | 25-49% | 48 (11.1) |
| | | 50-74% | 37 (8.6) |
| | | ≥75% | 27 (6.3) |
| | | *Don't know* | *53 (12.3)* |
| | | *Not applicable to my practice* | *185 (42.9)* |
| | Ablative procedures | 0% | 20 (4.6) |
| | | 1-24% | 42 (9.7) |
| | | 25-49% | 37 (8.6) |
| | | 50-74% | 37 (8.6) |
| | | ≥75% | 35 (8.1) |
| | | *Don't know* | *54 (12.5)* |
| | | *Not applicable to my practice* | *206 (47.8)* |
| | Hysterectomy | 0% | 21 (4.9) |
| | | 1-24% | 44 (10.2) |
| | | 25-49% | 45 (10.4) |
| | | 50-74% | 41 (9.5) |
| | | ≥75% | 27 (6.3) |
| | | *Don't know* | *59 (13.7)* |
| | | *Not applicable to my practice* | *194 (45.0)* |
| | Chemotherapy | 0% | 20 (4.6) |
| | | 1-24% | 39 (9.1) |
| | | 25-49% | 40 (9.3) |
| | | 50-74% | 25 (5.8) |
| | | ≥75% | 26 (6.0) |
| | | *Don't know* | *55 (12.8)* |
| | | *Not applicable to my practice* | *226 (52.4)* |
| | Radiation | 0% | 19 (4.4) |
| | | 1-24% | 32 (7.4) |
| | | 25-49% | 33 (7.7) |
| | | 50-74% | 37 (8.6) |
| | | ≥75% | 21 (4.9) |
| | | *Don't know* | *62 (14.4)* |

| | | | |
|---|---|---|---|
| | | *Not applicable to my practice* | *227 (52.7)* |
| | Cold knife conization | 1 week to <2 weeks | 13 (3.0) |
| | | 2 weeks to <4 weeks | 31 (7.2) |
| | | 1 month to <2 months | 32 (7.4) |
| | | 2 months to <4 months | 42 (9.7) |
| | | 4 months to <6 months | 33 (7.7) |
| | | >6 months | 13 (3.0) |
| | | *Don't know* | *48 (11.1)* |
| | | *Not applicable to my practice* | *219 (50.8)* |
| | Other excisional | 1 week to <2 weeks | 19 (4.4) |
| | | 2 weeks to <4 weeks | 32 (7.4) |
| | | 1 month to <2 months | 52 (12.1) |
| | | 2 months to <4 months | 33 (7.7) |
| | | 4 months to <6 months | 33 (7.7) |
| | | >6 months | 16 (3.7) |
| | | *Don't know* | *48 (11.1)* |
| | | *Not applicable to my practice* | *198 (45.9)* |
| Q42 Length of **deferral period** for **postponed treatment procedures** | Ablative procedures | 1 week to <2 weeks | 17 (3.9) |
| | | 2 weeks to <4 weeks | 18 (4.2) |
| | | 1 month to <2 months | 40 (9.3) |
| | | 2 months to <4 months | 42 (9.7) |
| | | 4 months to <6 months | 30 (7.0) |
| | | >6 months | 20 (4.6) |
| | | *Don't know* | *50 (11.6)* |
| | | *Not applicable to my practice* | *214 (49.7)* |
| | Hysterectomy | 1 week to <2 weeks | 21 (4.9) |
| | | 2 weeks to <4 weeks | 23 (5.3) |
| | | 1 month to <2 months | 31 (7.2) |
| | | 2 months to <4 months | 46 (10.7) |
| | | 4 months to <6 months | 30 (7.0) |
| | | >6 months | 20 (4.6) |
| | | *Don't know* | *55 (12.8)* |
| | | *Not applicable to my practice* | *205 (47.6)* |
| | Chemotherapy | 1 week to <2 weeks | 19 (4.4) |
| | | 2 weeks to <4 weeks | 28 (6.5) |
| | | 1 month to <2 months | 32 (7.4) |
| | | 2 months to <4 months | 27 (6.3) |
| | | 4 months to <6 months | 28 (6.5) |
| | | >6 months | 22 (5.1) |
| | | *Don't know* | *49 (11.4)* |
| | | *Not applicable to my practice* | *226 (52.4)* |
| | Radiation | 1 week to <2 weeks | 17 (3.9) |
| | | 2 weeks to <4 weeks | 28 (6.5) |
| | | 1 month to <2 months | 29 (6.7) |
| | | 2 months to <4 months | 36 (8.4) |
| | | 4 months to <6 months | 25 (5.8) |
| | | >6 months | 11 (2.6) |
| | | *Don't know* | *55 (12.8)* |
| | | *Not applicable to my practice* | *230 (53.4)* |

### Supplementary Figures

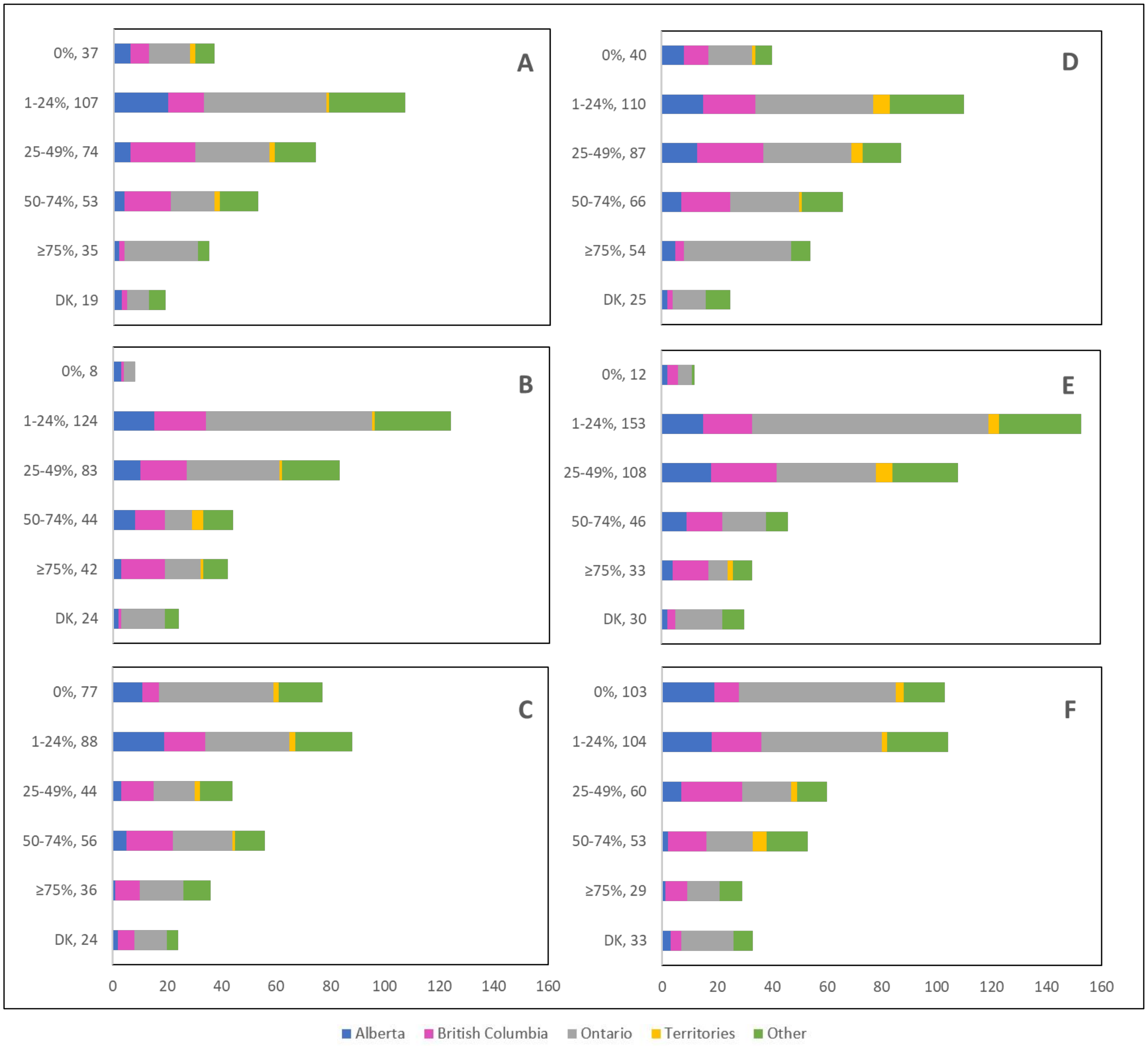


**Supplementary Figure 1:** Cancelled (n = 325) and postponed (n = 382) **screening** appointments by **province** that were cancelled or postponed by physician/providers' institution, by patient, or converted to telemedicine

**Supplementary Figure 1 legend**:
Number cancelled by (A) physician or providers' institution, (B) patient, and (C) converted to telemedicine.
Number postponed by (D) physician or providers' institution, (E) patient, and (F) converted to telemedicine.
Answers include responses for questions 7 (cancellations) and 9 (postponements) by question 2 (province). Respondents were asked to ensure that their answers did not exceed 100% for each question. (i.e., for each respondent, A + B + C ≈ 100% and D + E + F ≈ 100%).
The **x axis** represents frequency of responses by province. Territories include Northwest Territories, Nunavut, and Yukon. Other provinces include Manitoba, New Brunswick, Newfoundland and Labrador, Nova Scotia, Prince Edward Island, Quebec, and Saskatchewan (and one respondent who preferred not to say).
The **y axis** represents cancelled or postponed screening appointments using a predefined interval scale.
DK: Don't know

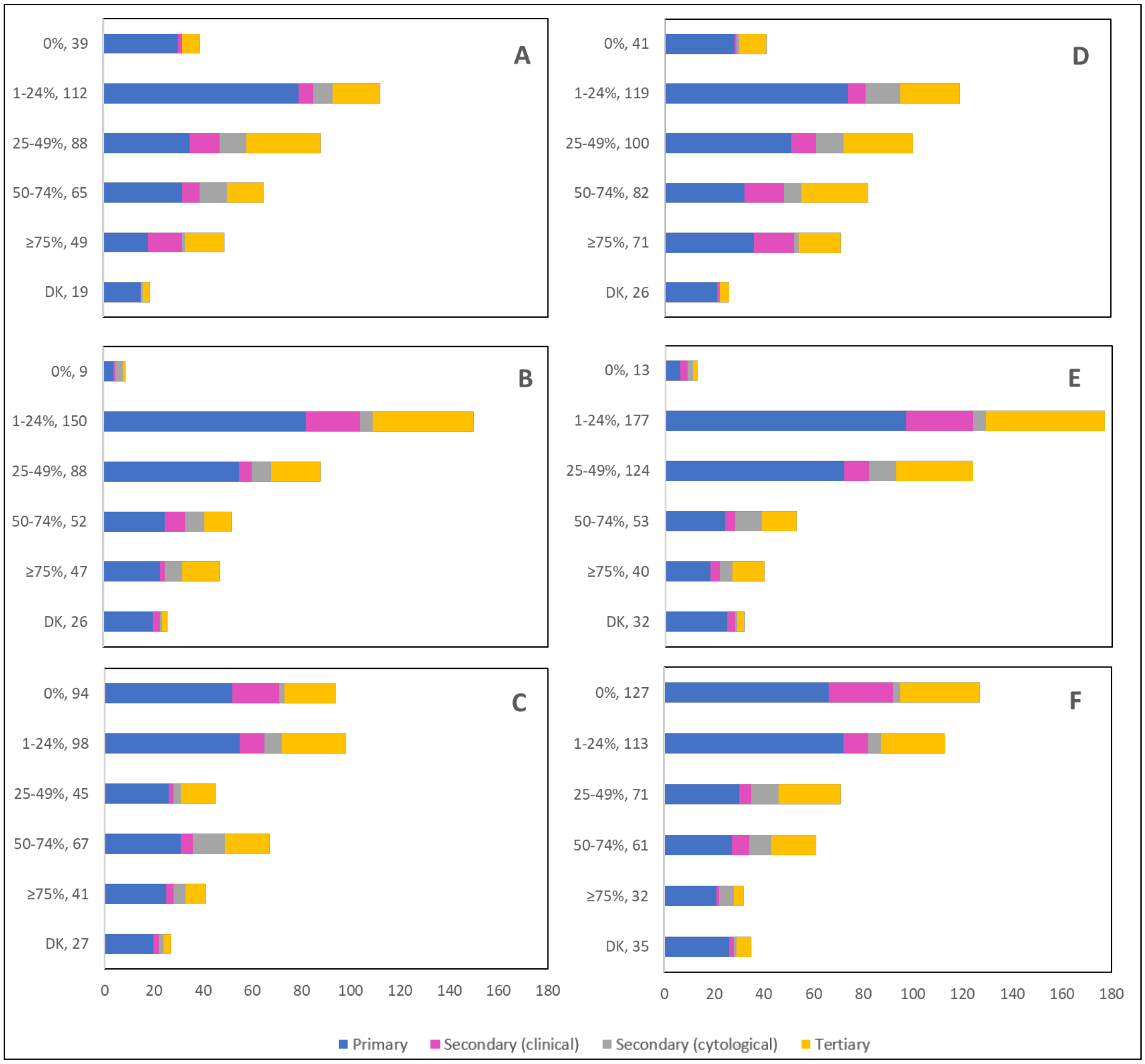


**Supplementary Figure 2:** Cancelled (n = 325) and postponed (n = 382) **screening** appointments by **profession** that were cancelled or postponed by physician/providers' institution, by patient, or converted to telemedicine

**Supplementary Figure 2 legend**:
Number cancelled by (A) physician or providers' institution, (B) patient, and (C) converted to telemedicine.
Number postponed by (D) physician or providers' institution, (E) patient, and (F) converted to telemedicine.
Answers include responses for questions 7 (cancellations) and 9 (postponements) by question 4 (profession).
Respondents were asked to ensure that their answers did not exceed 100% for each question. (i.e., for each respondent, A + B + C ≈ 100% and D + E + F ≈ 100%).
The **x axis** represents frequency of responses by profession. Primary includes general practitioners/family physicians, nurse practitioners/registered nurses, physician assistants and a manager of a community hospital; Secondary (clinical) includes colposcopists and colposcopy registered nurses/registered practical nurses; Secondary (cytological) includes cytopathologists/technologists and pathologists; Tertiary includes gynecologists/obstetrician-gynecologists, gynecology oncologists, and gynecology nurses. Frequency count exceeded total number of respondents as some reported multiple professions.
The **y axis** represents cancelled or postponed screening appointments using a predefined interval scale.
DK: Don't know

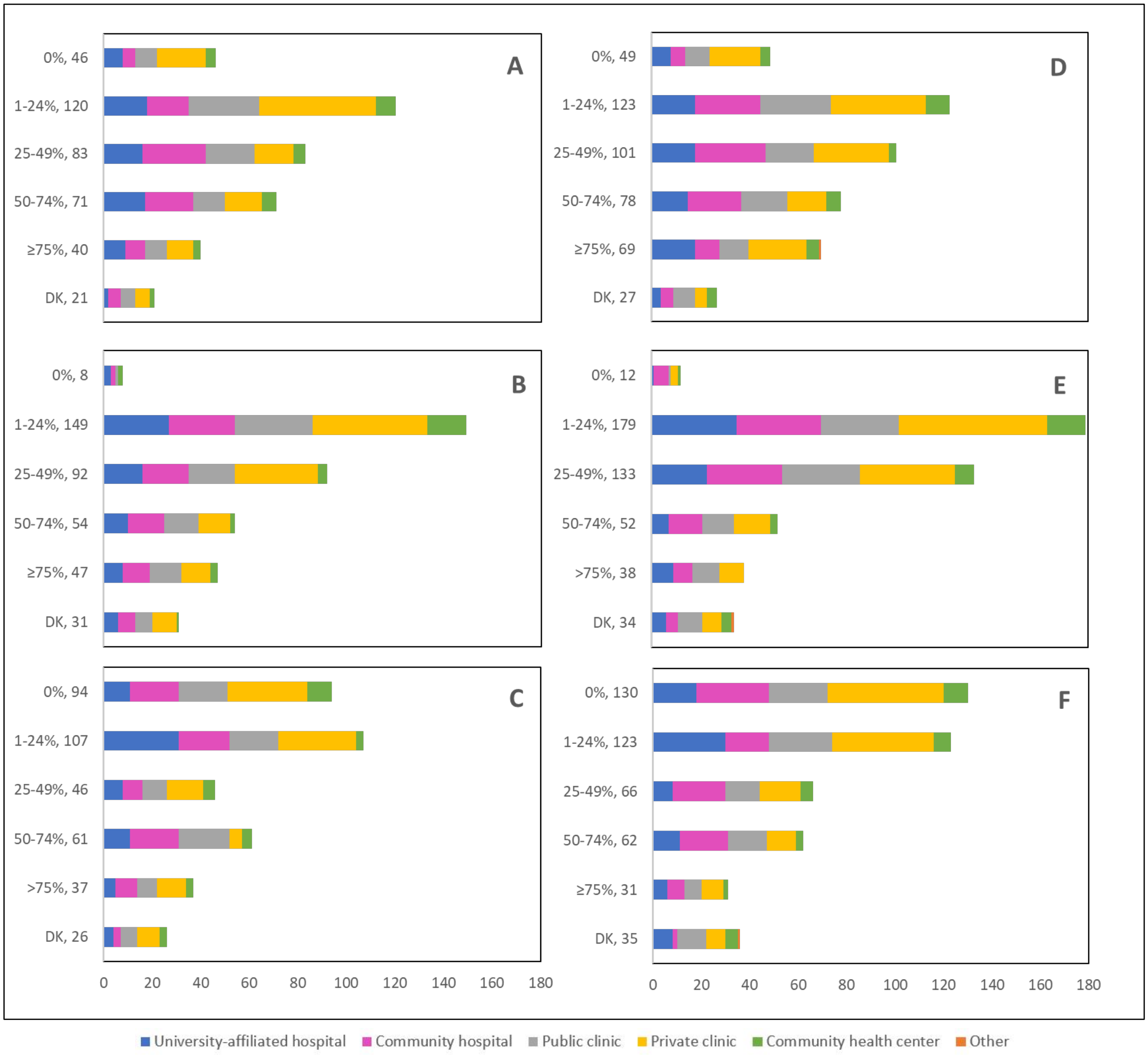


**Supplementary Figure 3:** Cancelled (n = 325) and postponed (n = 382) **screening** appointments by **place of practice** that were cancelled or postponed by physician/providers' institution, by patient, or converted to telemedicine

**Supplementary Figure 3 legend**:
Number cancelled by (A) physician or providers' institution, (B) patient, and (C) converted to telemedicine.
Number postponed by (D) physician or providers' institution, (E) patient, and (F) converted to telemedicine.
Answers include responses for questions 7 (cancellations) and 9 (postponements) by question 5 (place of practice).
Respondents were asked to ensure that their answers did not exceed 100% for each question. (i.e., for each respondent, A + B + C ≈ 100% and D + E + F ≈ 100%).
The **x axis** represents frequency of responses by place of practice. Frequency count exceeded total number of respondents as some reported multiple places of practice.
The **y axis** represents cancelled or postponed screening appointments using a predefined interval scale.
DK: Don't know

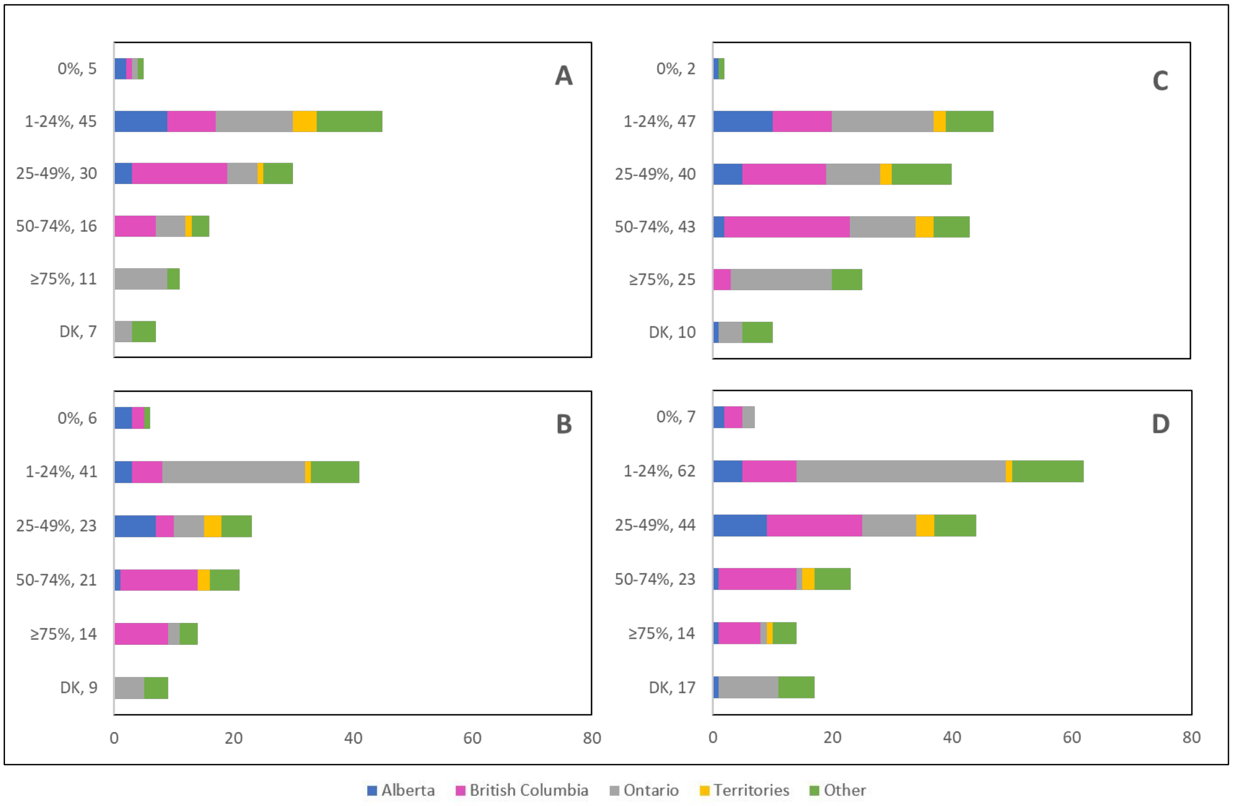


**Supplementary Figure 4:** Cancelled (n = 114) and postponed (n = 167) **colposcopy** appointments by **province** that were cancelled or postponed by physician/providers' institution or by patient

**Supplementary Figure 4 legend**:
Number cancelled by (A) physician or providers' institution and (B) patient.
Number postponed by (C) physician or providers' institution and (D) patient.
Answers include responses for questions 26 (cancellations) and 28 (postponements) by question 2 (province).
Respondents were asked to ensure that their answers did not exceed 100% for each question. (i.e., for each respondent, A + B ≈ 100% and C + D ≈ 100%).
The **x axis** represents frequency of responses by province. Territories include Northwest Territories, Nunavut, and Yukon. Other provinces include Manitoba, New Brunswick, Newfoundland and Labrador, Nova Scotia, Prince Edward Island, Quebec, and Saskatchewan (and one respondent who preferred not to say).
The **y axis** represents cancelled or postponed colposcopy appointments using a predefined interval scale.
DK: Don't know

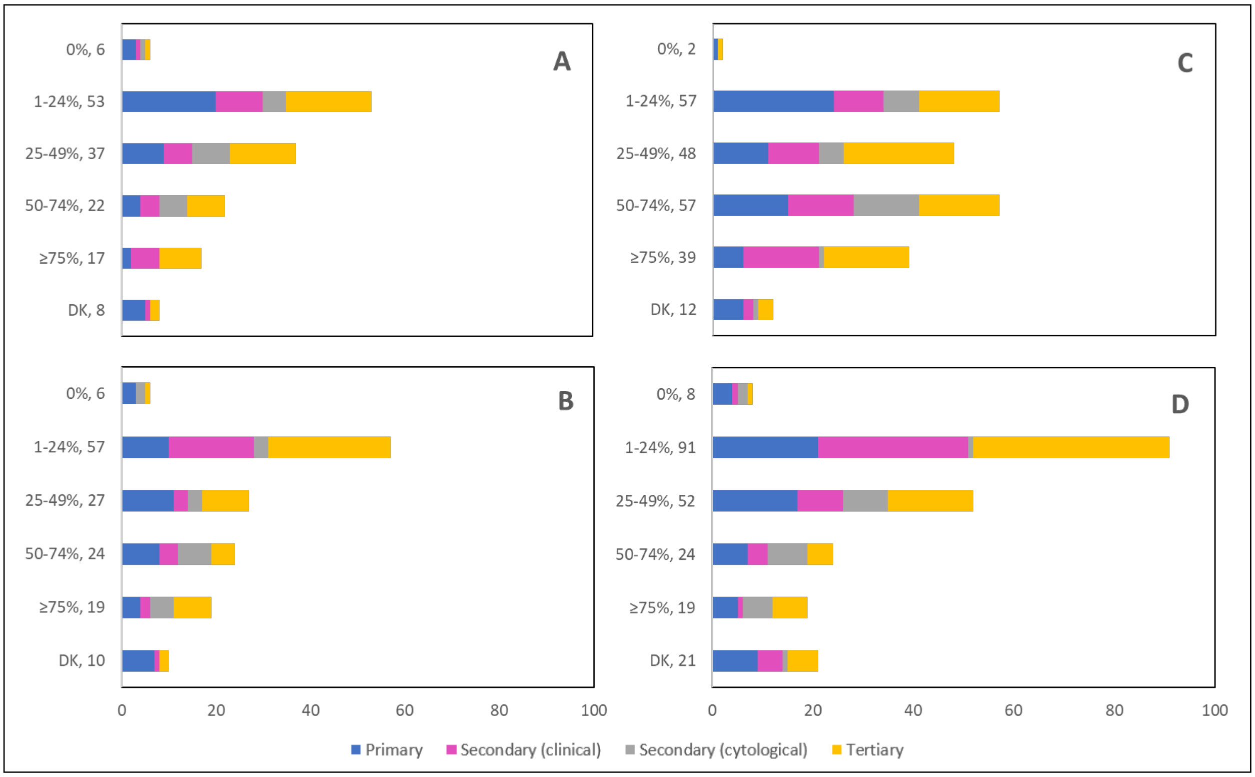


**Supplementary Figure 5:** Cancelled (n = 114) and postponed (n = 167) **colposcopy** appointments by **profession** that were cancelled or postponed by physician/providers' institution or by patient

**Supplementary Figure 5 legend**:
Number cancelled by (A) physician or providers' institution and (B) patient.
Number postponed by (C) physician or providers' institution and (D) patient.
Answers include responses for questions 26 (cancellations) and 28 (postponements) by question 4 (profession).
Respondents were asked to ensure that their answers did not exceed 100% for each question. (i.e., for each respondent, A + B ≈ 100% and C + D ≈ 100%).
The **x axis** represents frequency of responses by profession. Primary includes general practitioners/family physicians, nurse practitioners/registered nurses, and physician assistants; Secondary (clinical) includes colposcopists and colposcopy registered nurses/registered practical nurses; Secondary (cytological) includes cytopathologists/technologists and pathologists; Tertiary includes gynecologists/obstetrician-gynecologists, gynecology oncologists, and gynecology nurses. Frequency count exceeded total number of respondents as some reported multiple professions.
The **y axis** represents cancelled or postponed colposcopy appointments using a predefined interval scale.
DK: Don't know

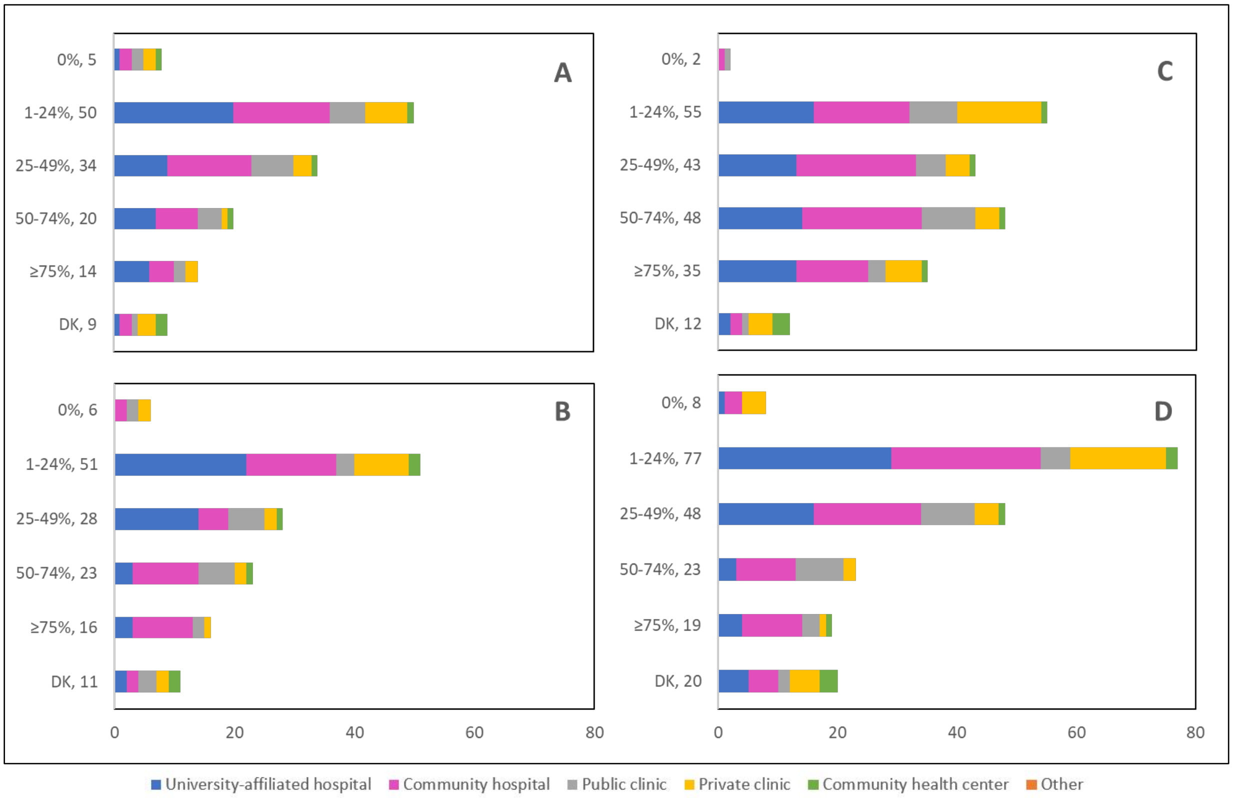


**Supplementary Figure 6:** Cancelled (n = 114) and postponed (n = 167) **colposcopy** appointments by **place of practice** that were cancelled or postponed by physician/providers' institution or by patient

**Supplementary Figure 6 legend**:
Number cancelled by (A) physician or providers' institution and (B) patient.
Number postponed by (C) physician or providers' institution and (D) patient.
Answers include responses for questions 26 (cancellations) and 28 (postponements) by question 5 (place of practice). Respondents were asked to ensure that their answers did not exceed 100% for each question. (i.e., for each respondent, A + B ≈ 100% and C + D ≈ 100%).
The **x axis** represents frequency of responses by place of practice. Frequency count exceeded total number of respondents as some reported multiple places of practice.
The **y axis** represents cancelled or postponed colposcopy appointments using a predefined interval scale.
DK: Don't know

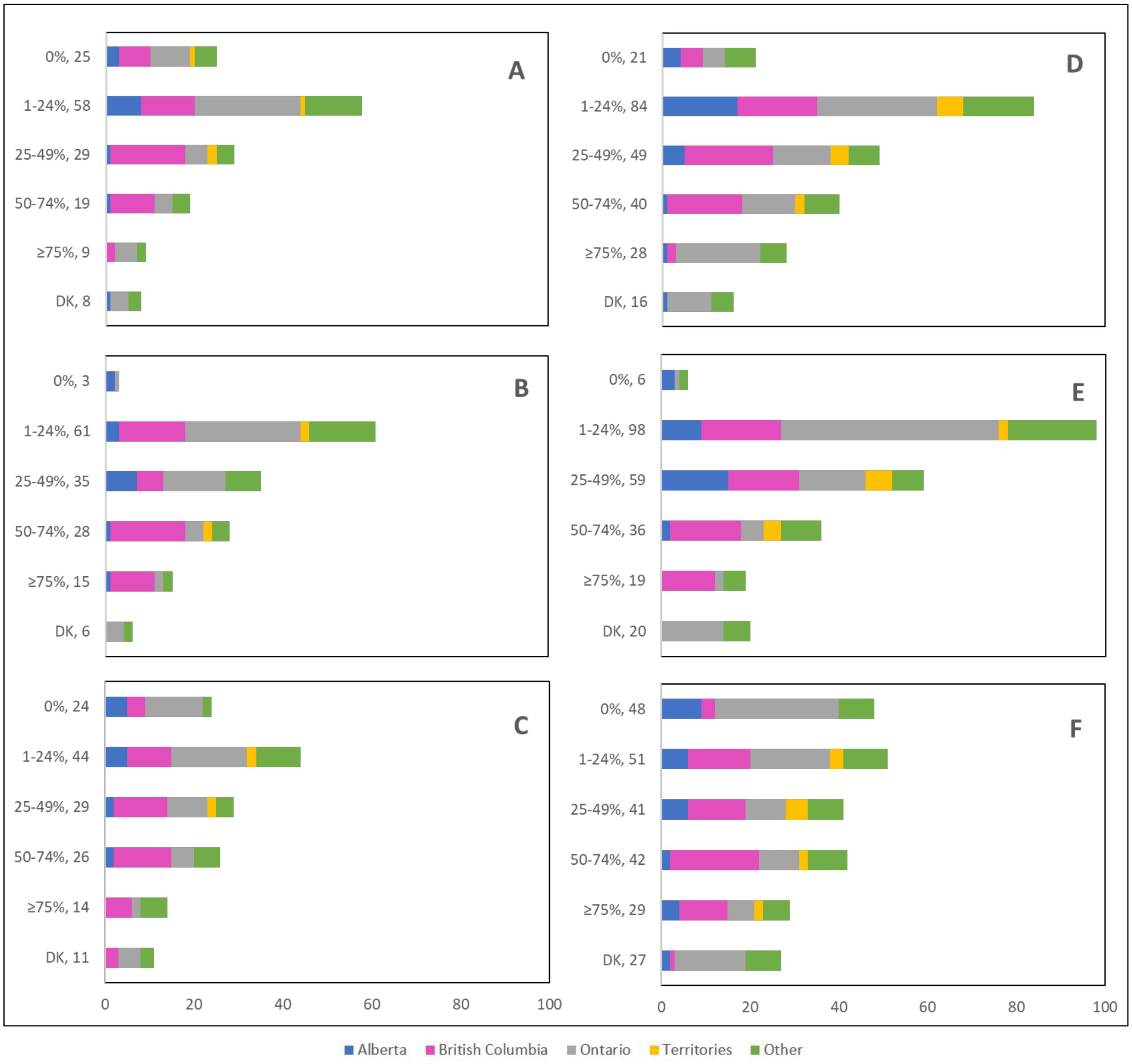


**Supplementary Figure 7:** Cancelled (n = 148) and postponements (n = 238) of **follow-up** appointments by **province** that were cancelled or postponed by physician/providers' institution, by patient, or converted to telemedicine

**Supplementary Figure 7 legend**:
Number cancelled by (A) physician or providers' institution, (B) patient and (C) converted to telemedicine.
Number postponed by (D) physician or providers' institution, (E) patient and (F) converted to telemedicine.
Answers include responses for questions 32 (cancellations) and 34 (postponements) by question 2 (province).
Respondents were asked to ensure that their answers did not exceed 100% for each question. (i.e., for each respondent, A + B + C ≈ 100% and D + E + F ≈ 100%).
The **x axis** represents frequency of responses by province. Territories include Northwest Territories, Nunavut, and Yukon. Other provinces include Manitoba, New Brunswick, Newfoundland and Labrador, Nova Scotia, Prince Edward Island, Quebec, and Saskatchewan (and one respondent who preferred not to say).
The **y axis** represents cancelled or postponed follow-up appointments using a predefined interval scale.
DK: Don't know

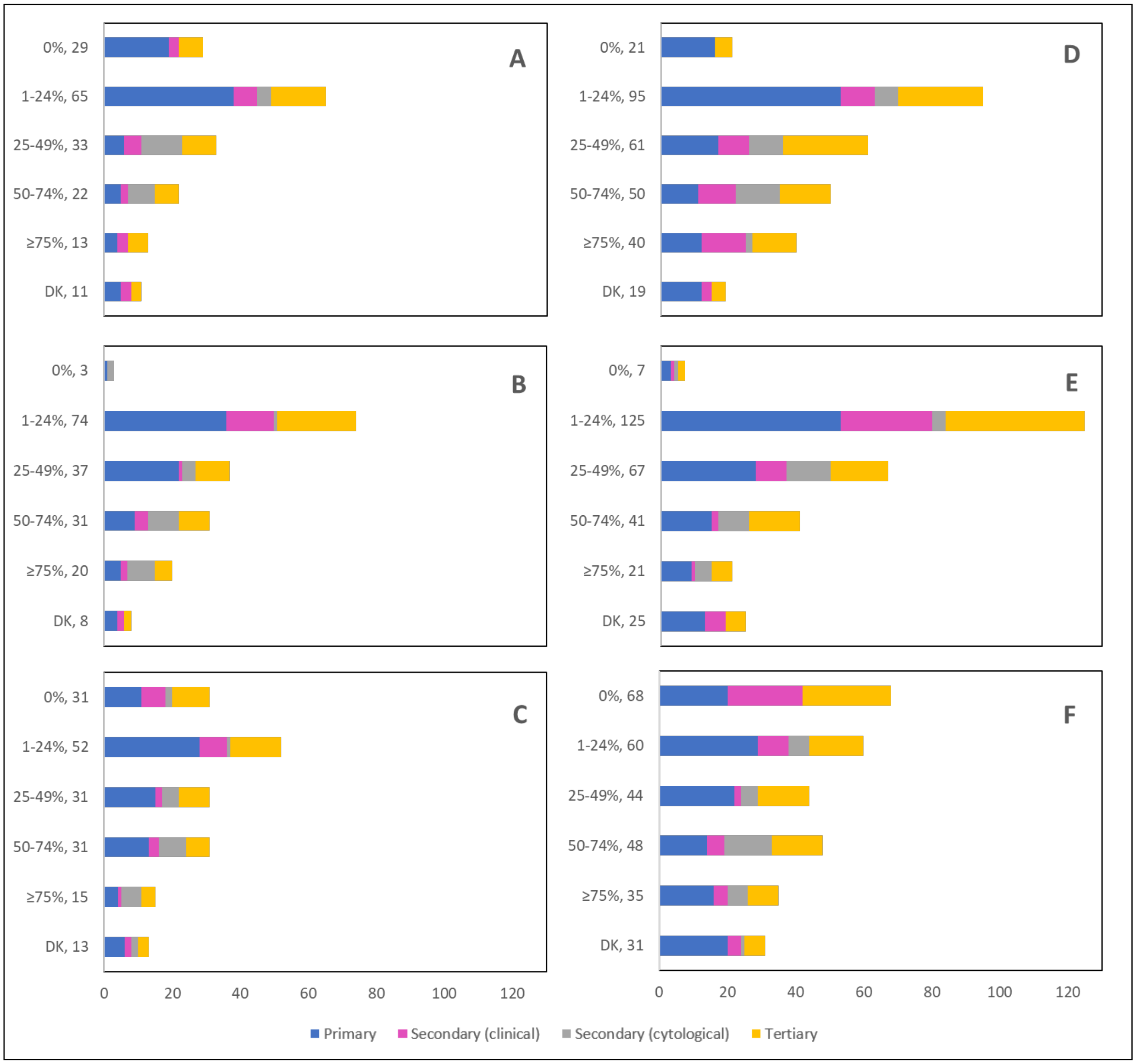


**Supplementary Figure 8:** Cancelled (n = 148) and postponed (n = 238) **follow-up** appointments by **profession** that were cancelled or postponed by physician/providers' institution, by patient, or converted to telemedicine

**Supplementary Figure 5 legend**:
Number cancelled by (A) physician or providers' institution, (B) patient and (C) converted to telemedicine.
Number postponed by (D) physician or providers' institution, (E) patient and (F) converted to telemedicine.
Answers include responses for questions 32 (cancellations) and 34 (postponements) by question 4 (profession).
Respondents were asked to ensure that their answers did not exceed 100% for each question. (i.e., for each respondent, A + B + C ≈ 100% and D + E + F ≈ 100%).
The **x axis** represents frequency of responses by profession. Primary includes general practitioners/family physicians, nurse practitioners/registered nurses, and physician assistants; Secondary (clinical) includes colposcopists and colposcopy registered nurses/registered practical nurses; Secondary (cytological) includes cytopathologists/technologists and pathologists; Tertiary includes gynecologists/obstetrician-gynecologists, gynecology oncologists, and gynecology nurses. Frequency count exceeded total number of respondents as some reported multiple professions.
The **y axis** represents cancelled or postponed follow-up appointments using a predefined interval scale.
DK: Don't know

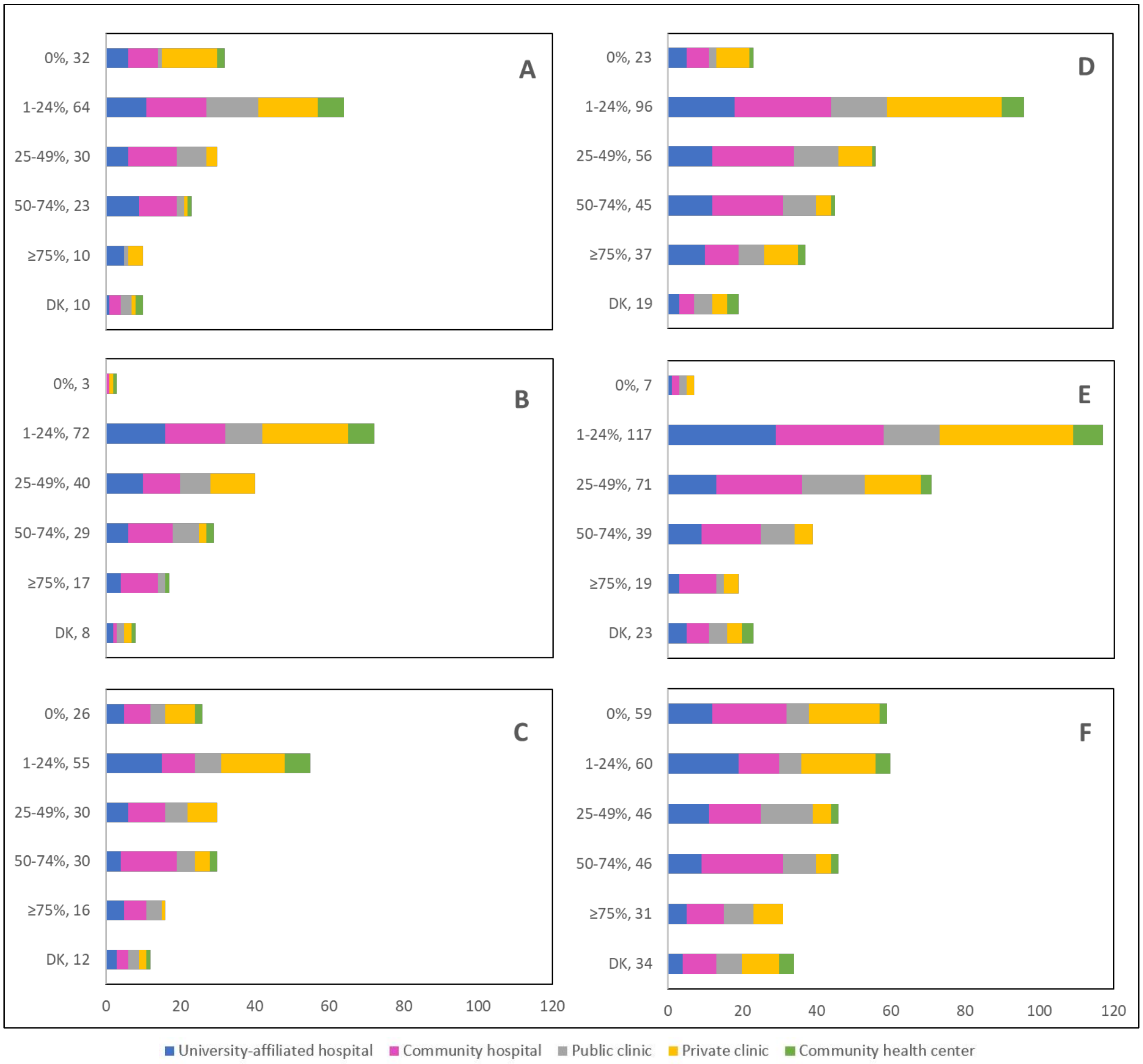


**Supplementary Figure 9:** Cancelled (n = 148) and postponed (n = 238) **follow-up** appointments by **place of practice** that were cancelled or postponed by physician/providers' institution, by patient, or converted to telemedicine

**Supplementary Figure 9 legend**:
Number cancelled by (A) physician or providers' institution, (B) patient and (C) converted to telemedicine.
Number postponed by (C) physician or providers' institution, (D) patient and (E) converted to telemedicine.
Answers include responses for questions 32 (cancellations) and 34 (postponements) by question 5 (place of practice).
Respondents were asked to ensure that their answers did not exceed 100% for each question. (i.e., for each respondent, A + B + C ≈ 100% and D + E + F ≈ 100%).
The **x axis** represents frequency of responses by place of practice. Frequency count exceeded total number of respondents as some reported multiple places of practice.
The **y axis** represents cancelled or postponed follow-up appointments using a predefined interval scale.
DK: Don't know

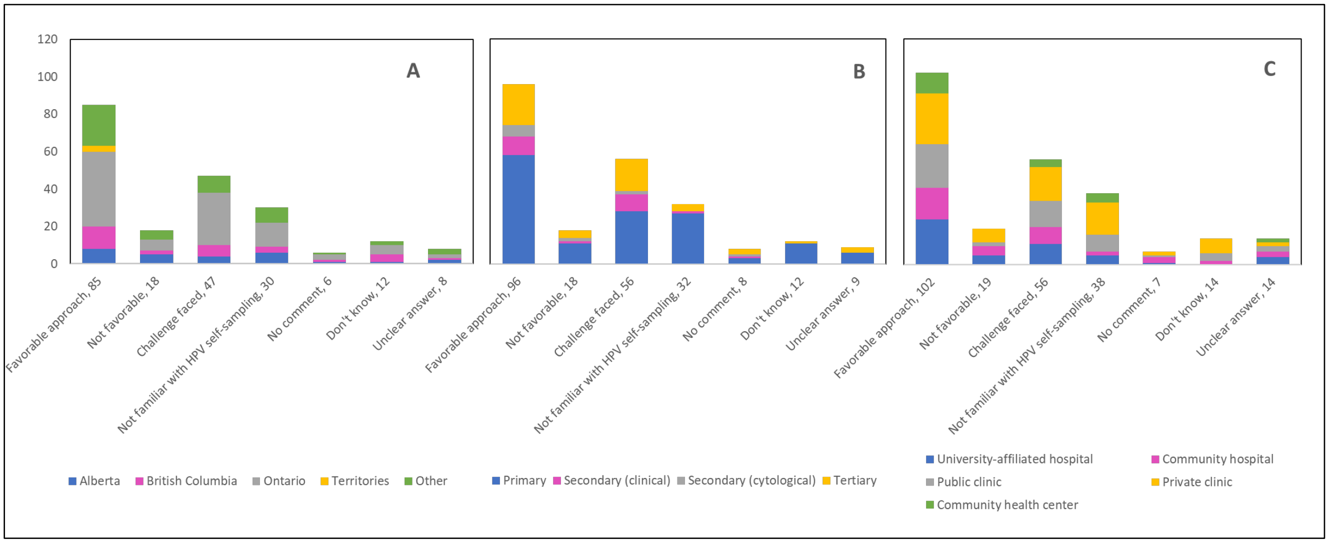


**Supplementary Figure 10:** Responses to **open-ended Q22** by province, profession and place of practice (n = 206)

**Supplementary Figure 10 legend:**
Opinions and perspectives are shown by (A) province, (B) profession, and (C) place of practice.
Answers include responses for question 22 (COVID-19 to encourage/facilitate/accelerate implementation of HPV self-sampling) by questions 2 (province), 4 (profession) and 5 (place of practice).
**Panel A:** Territories include Northwest Territories, Nunavut, and Yukon. Other provinces include Manitoba, New Brunswick, Newfoundland and Labrador, Nova Scotia, Prince Edward Island, Quebec, and Saskatchewan (and one respondent who preferred not to say).
**Panel B:** Primary includes general practitioners/family physicians, nurse practitioners/registered nurses, physician assistants and a manager of a community center; Secondary (clinical) includes colposcopists and colposcopy registered nurses/registered practical nurses; Secondary (cytological) includes cytopathologists/technologists and pathologists; Tertiary includes gynecologists/obstetrician-gynecologists, gynecology oncologists, and gynecology nurses.
**Panels B andC:** Frequency count exceeded total number of respondents as some reported multiple places of practice.

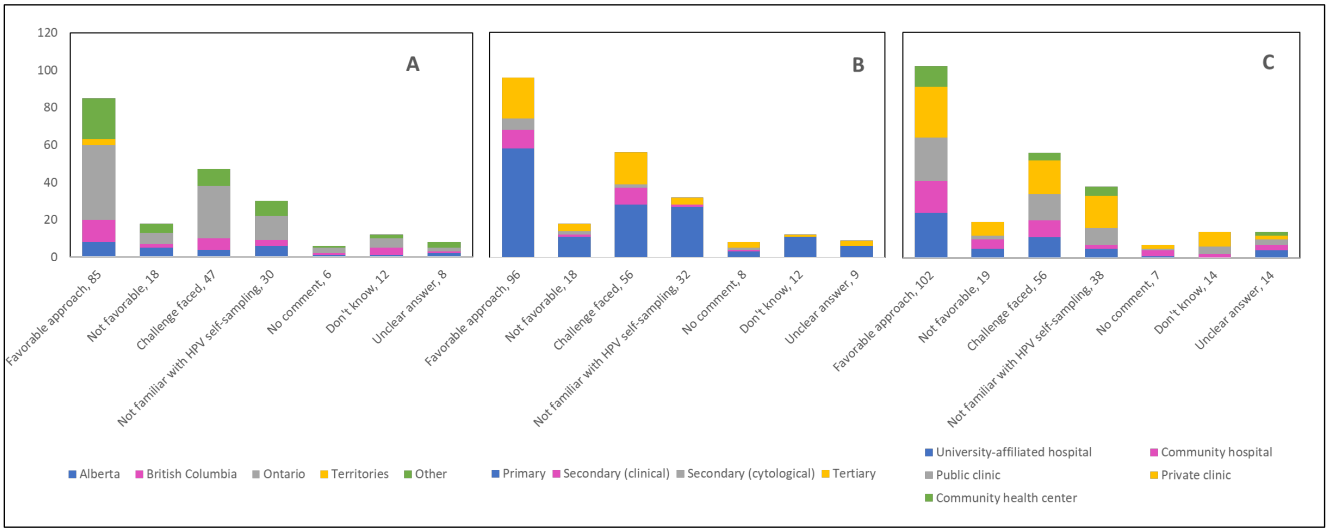


**Supplementary Figure 11:** Responses to **open-ended Q23** by province, profession and place of practice (n = 197)

**Supplementary Figure 11 legend:**
Opinions and perspectives are shown by (A) province, (B) profession, and (C) place of practice.
Answers include responses for question 23 (in favor of implementing HPV self-sampling as alternative screening method) by questions 2 (province), 4 (profession) and 5 (place of practice).
**Panel A:** Territories include Northwest Territories, Nunavut, and Yukon. Other provinces include Manitoba, New Brunswick, Newfoundland and Labrador, Nova Scotia, Prince Edward Island, Quebec, and Saskatchewan (and one respondent who preferred not to say).
**Panel B:** Primary includes general practitioners/family physicians, nurse practitioners/registered nurses, physician assistants, and a manager of a community health center; Secondary (clinical) includes colposcopists and colposcopy registered nurses/registered practical nurses; Secondary (cytological) includes cytopathologists/technologists and pathologists; Tertiary includes gynecologists/obstetrician-gynecologists, gynecology oncologists, and gynecology nurses.
**Panels B and C:** Frequency count exceeded total number of respondents as some reported multiple places of practice.
